# The Effects of Daylight Saving Time Clock Changes on Mental and Physical Health in England: Evidence from the Clinical Practice Research Datalink (CPRD)

**DOI:** 10.1101/2025.05.14.25327580

**Authors:** Melanie A de Lange, Kate Birnie, Rebecca C Richmond, Chin Yang Shapland, Sophie V Eastwood, Kate Tilling, Neil M Davies

## Abstract

**OBJECTIVES:** To explore the effects of daylight saving time clock changes on mental and physical health events in primary and secondary care in England.

**DESIGN:** Population-based study using linked electronic health records.

**SETTING:** English primary care practices contributing to the Clinical Practice Research Datalink (CPRD) Gold database, linked to hospital admissions and Accident and Emergency (A&E) data.

**PARTICIPANTS:** 683,809 individuals (road traffic injuries: all ages, cardiovascular disease (CVD): aged ≥40 years, all other conditions: ≥10 years) registered with a participating English GP practice, with a health event for one of our health conditions in their primary or secondary care record in the eight weeks surrounding the Spring or Autumn clock changes between 2008 and 2019.

**MAIN OUTCOME MEASURES:** Health events were defined as a diagnosis code (or symptom code and prescription for mental health outcomes in primary care) of anxiety, major acute CVD, depression, eating disorder, road traffic injury, self-harm or sleep disorder in primary or secondary care, or psychiatric condition in A&E. Negative binomial regression models, adjusted for day of the week and region (and Easter weekend in Spring), compared event rates in the week after the clock changes to the control period (four weeks before the changes and weeks two-four after).

**RESULTS:** In the week after the Autumn clock change five health conditions had fewer events, including: anxiety (IRR 0.97, 95% CI 0.95 to 0.98), acute CVD (IRR 0.98, 95% CI 0.96 to 0.999), depression (IRR 0.96, 95% CI 0.95 to 0.97), psychiatric conditions (IRR: 0.94, 95% CI 0.90 to 0.98) and sleep disorders (IRR 0.92, 95% CI 0.87 to 0.97). There was little evidence of reductions in eating disorder diagnoses, road traffic injuries or self-harm.

**CONCLUSIONS:** There was a reduction in rates of CVD, sleep disorders and mental health disorders after the Autumn clock change, but little evidence that the Spring clock change affected health.

**What is already known on this topic:**

- Studies suggest that the DST clock changes, particularly the Spring clock change, have a detrimental effect on population health.
- On this basis, sleep societies have unanimously called for the clock changes to be abolished and recommend GMT be adopted year-round.
- In 2019, the EU voted to end the practice of changing the clocks. However, it is not clear whether England will do the same.

**What this study adds:**

- This extensive study of the effects of the DST clock changes on health in England used a large, representative GP and hospital dataset, and examined multiple health conditions.
- This study estimates the effects of the clock changes on demand for NHS services.
- In the week after the Autumn clock change there was a decrease in health events for sleep disorders, CVD, anxiety, depression and psychiatric conditions.

## Introduction

Daylight saving time (DST) was introduced during World War One and involves moving the clocks one hour forward in Spring and back one hour in Autumn. DST operates in around 70 countries^1^ and affects a quarter of the world’s population^2^. However, in recent years the clock changes have become the focus of intense debate.

Research suggests that the clock changes may be detrimental to population health. For example, a meta-analysis of 12 studies from ten countries reported that the risk of acute myocardial infarction (AMI) increased by 4% in the week after the Spring clock change.^3^ Furthermore, a study using 20 years of US registry data identified a 6% rise in fatal traffic accidents in the week after the Spring clock change^4^ and an analysis of nationwide psychiatric hospitals in Denmark found an 11% rise in the incidence rate of unipolar depression after the Autumn clock change.^5^ However, other studies have found little evidence that the clock changes increase the risk of adverse health events.^6–13^ Notably, we have little evidence from England and studies have not estimated the effect of the clock changes on the use of primary care services.

In the last decade, countries such as the USA^14^, Mexico^15^, Turkey^16^ and Jordan^17^ have ended or moved towards ending the clock changes. In 2019, the European Union voted to do the same^18^, although this policy has yet to be implemented. The EU’s decision did not prompt an immediate policy change in the UK. However, a subsequent House of Lords inquiry highlighted the need for more research.^19^ In late 2024, the British Sleep Society joined the appeal from other sleep societies for the DST clock changes to be abolished and recommended that permanent standard time (Greenwich Mean Time) be adhered to throughout the year.^20^ In March 2025, in a debate over the possible benefits of implementing double British Summer Time (having the clocks two hours ahead of GMT in summer and one hour ahead in winter), the UK government stated that the current evidence for altering the existing DST system was not overwhelming but raised some interesting issues, such as the effects of the clock changes on road safety and mental health^21^.

Here, we used a large database of linked primary and secondary care records to investigate the effects of the clock changes on people’s health in England. The health conditions we examined were anxiety, major acute CVD, depression, eating disorder, road traffic injury, self-harm or sleep disorders in primary or secondary care (hospital admissions and A&E), as well as psychiatric conditions in A&E.

## Methods

### Data Source

This research was pre-specified in CPRD protocol 22/002468 (see supplementary text S1 for protocol and S2 for minor deviations from the protocol). We used the Clinical Practice Research Datalink (CPRD) GOLD database. This is a UK longitudinal primary care database established in 1987. It contains anonymised, routinely collected medical records from participating primary care practices using the Vision® software system.^22^ The July 2024 version used in this study contained the records of over 21.5 million people from 985 primary care practices. As of mid-2024, 4.3% of the UK population and 4.4% of UK primary care practices actively contributed to the database.^23^

CRPD GOLD is broadly representative of the UK population in terms of age, sex, and ethnicity compared with the UK census in 2011.^22^ Data include diagnoses, symptoms, prescriptions, tests, vaccinations, referrals, demographics and lifestyle information.^22^ We also used linked data on hospital admissions from Hospital Episode Statistics Admitted Patient Care (HES APC), hospital accident and emergency visits (HES A&E) and Office for National Statistics (ONS) Index of Multiple Deprivation (IMD) (based on patient postcode). Because linked data were only available for 75% of English practices^22^, our study was restricted to England.

### Study Population

To be included in this study, patients had to be registered at an English GP practice, have a record classified as ‘acceptable’ by CPRD, be registered at an ‘up to standard’ GP practice (in terms of practice data quality) and be eligible for data linkage to HES APC, HES A&E and Patient Level IMD. Patients also had to have at least 1 year of research-quality follow-up time in CPRD GOLD during the study period (established by CPRD using practice and patient-level data quality checks). We also required patients to have a health event (defined below) for one of our eight health conditions of interest in their primary or secondary care record within the 8 weeks surrounding the Spring or Autumn clock changes between 2008 and 2019. Because this study looks at the numbers of events in particular 8-week periods, and we can assume that the population remains fairly constant over these small periods of time, only people with events contributed to the analysis. For quality control purposes, we excluded primary care events outside of a patient’s valid GP registration period (based on practice and patient-level data quality checks). There were no age restrictions placed on road traffic injury events. However, for acute cardiovascular disease events to be included, patients had to be aged ≥ 40 years at the time of the clock change. For all other health conditions, patients had to be aged ≥10 years at the time of the clock change. Patients could have events for more than one of our health conditions during the study period and could contribute to both Spring and Autumn analyses. Our final sample consisted of 683,809 patients and 1,565,032 health events (see Figure 1).

**Figure 1.**
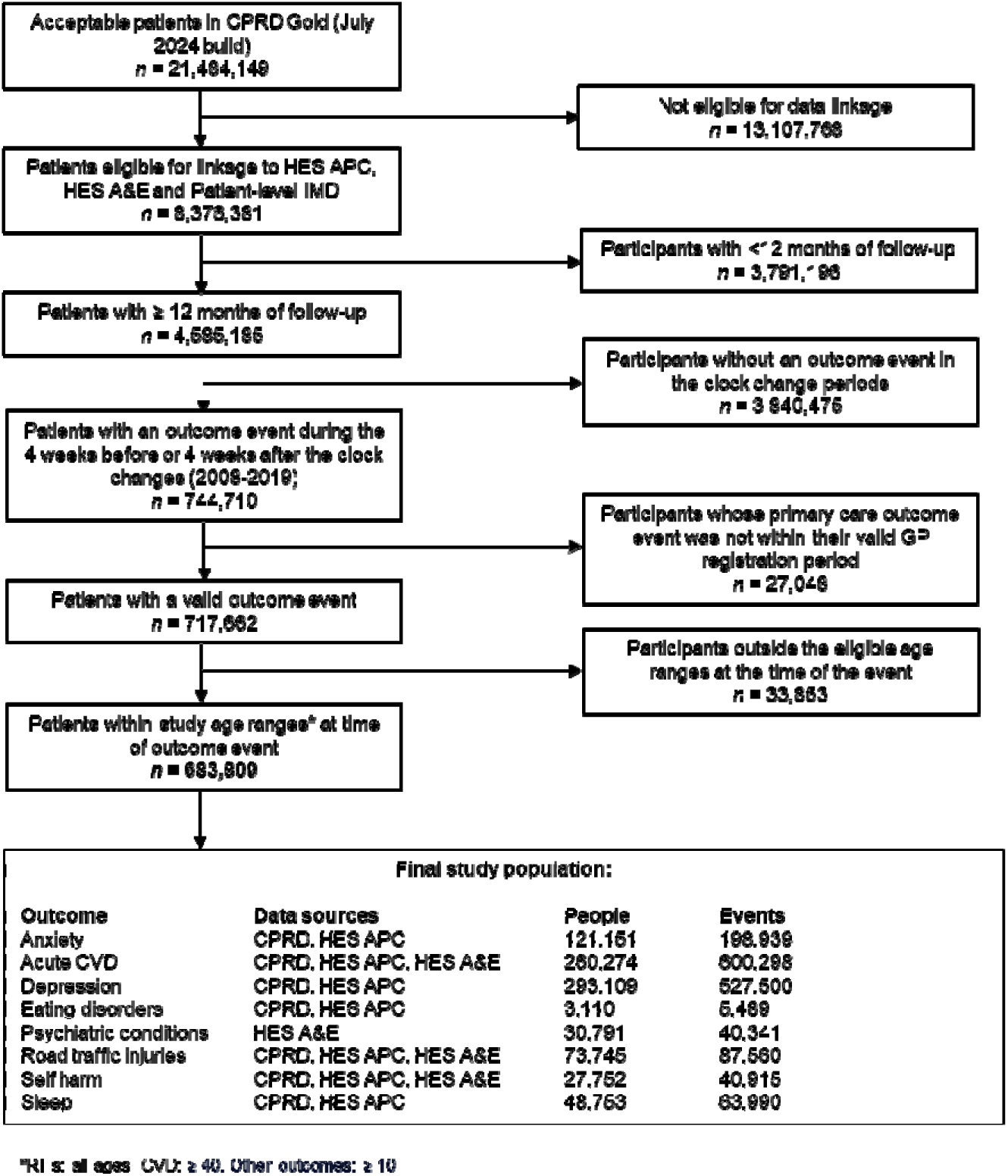
Participant flow diagram.

### Exposures

The exposures of interest were the Spring and Autumn clock changes between 2008 and 2019. In England, the clocks went forward one hour at 1am on the last Sunday and March and went back one hour at 2am on the last Sunday in October. A complete list of these dates is provided in supplementary table S1.

### Outcomes

Our eight health conditions of interest were anxiety, acute major CVD, depression, eating disorders, psychiatric conditions (A&E only), road traffic injuries, self-harm and sleep disorders (see Figure 1). We chose these conditions because previous research suggests that the clock changes affect CVD^3^ and road traffic injuries.^4^ The clock changes are also reported to affect sleep duration, efficiency (proportion of time in bed spent asleep) and fragmentation (number of awakenings).^24, 25^ It therefore seemed logical to examine the effects of the clock changes on sleep disorders, particularly given that our earlier study found electronic health records to be a valuable data source for measuring insomnia prevalence^26^. We also wanted to extend the scope of the existing literature on the effect of the clock changes on depression^5^, suicide^27^ and mental health visits to emergency departments^28^ to explore other mental health conditions that may be better captured in primary care data. We therefore included self-harm (including suicide), eating disorders and anxiety in our study. We were able to identify CVD, road traffic injuries and self-harm in all three healthcare datasets: primary care, HES APC and HES A&E data. However, A&E data is not specific enough to identify individual conditions such as depression, anxiety, sleep disorders or eating disorders. We, therefore, included psychiatric conditions in A&E data separately.

We defined health events in secondary care as the presence of a diagnosis code in a person’s health record that was entered during a hospital admission (HES APC data) or A&E consultation (HES A&E data). In HES APC data these were ICD-10 codes. The HES A&E dataset has its own coding system. We identified road traffic injury and self-harm events using the A&E patient group code, which gives the reason for the A&E episode. We identified CVD and psychiatric conditions events based on the patient group code combined with a two-digit diagnosis code (see supplementary text S3). We defined health events in primary care as the presence of a medical code recorded during a GP consultation.

For most of our health conditions these medical codes represented diagnoses. However, for sleep disorders, anxiety and depression, medical codes often relate to symptoms rather than diagnoses. For these conditions we counted medical codes as a health event if they were specific enough to identify the condition. However, where the codes related to symptoms that were not considered adequate to define the health condition, the health event was only counted if the patient also had a relevant prescription within the 90 day-period either side of the symptom code. Detailed definitions of all outcomes are provided in supplementary text S3. We included both incident and subsequent events for all health conditions.

Where possible, we created potential code lists based on existing, published lists (see https://github.com/MeldeLange/dst_cprd/code_lists for details). Details of how we created medical code and ICD-10 code lists for outcomes when no existing code lists were available are provided in supplementary text S4. All preliminary code lists were checked by a general practitioner (S.V.E) with experience using the codes in research and clinical practice. Based on their feedback, we refined the lists to ensure only relevant codes were included. The final code lists used in the analyses can be found at https://github.com/MeldeLange/dst_cprd/code_lists.

### Covariates

Our analysis was restricted to a short period of time either side of the clock change. This meant that we could assume there were no confounders of the effect of the clock changes on our outcomes. We considered several sociodemographic and lifestyle characteristics to be potential effect modifiers of the effect of the clock changes on health. We defined age and incident versus prevalent cases at the time of the clock change (exposure). Index of multiple deprivation (IMD) data were from 2019. Sex was recorded only once in the primary care data. We defined body mass index (BMI), alcohol status, smoking status, and systolic and diastolic blood pressure at the most recent record before the clock change. We identified CVD event subgroups based on disease codes in primary care and HES APC data. It was impossible to create subgroups for other outcomes or in A&E data because the codes used were not sufficiently specific. Full details of how we derived all covariates are provided in supplementary table S2. Briefly, they consisted of two categories plus a category for missing values (if necessary). We chose different age cut-offs for the eight health outcomes so that we had equally-sized binary categories based on the median value. Geographical region and year were considered to be potential effect modifiers. Region consisted of nine categories based on the Strategic Health Authority of each GP practice.

Other covariates that could effect the outcome only included the Easter weekend in Spring and day of the week. The Easter weekend and day of the week were considered to be competing exposures that may coincide with the clock change and could affect health outcomes, but are not related to the clock change. Dates of the 5-day Easter weekends in our study period are provided in supplementary table S3.

### Statistical analysis

#### Primary analysis

We plotted the total number of events per day for each health outcome for the eight-week period surrounding the Spring and Autumn clock changes. We conducted negative binomial regression analyses, adjusted for the day of the week and region (and Easter weekend in Spring), to estimate incidence rate ratios (IRR) and 95% confidence intervals (CIs) comparing the number of events in the first week after the clock changes to the average weekly number of events in the control period, which consisted of the four weeks before the clock changes and weeks two-four after the clock changes (See supplementary figure S1a). We chose a one-week exposure period because the effects of the clock changes on sleep are believed to last around a week^24^. It also meant that our results were comparable to the majority of studies examining the acute effects of the clock changes, which have focused on the first one or two weeks post-clock change^3, 9^. Our control period was chosen so that we had a relatively narrow window around the clock change to reduce the risk of other time-varying factors, such as other exposures or changes in the underlying population, biasing the results^29^. We also wanted to include weeks before and after the clock change in our control period so that we were not just capturing seasonal trends. We used negative binomial regression rather than Poisson regression to address overdispersion in the data. In our analyses we split our data by year (12 years), region (9 regions) and day (56 days in 8 week period), which gave us 6,048 data points in our primary analysis.

We adjusted the number of events per day to account for the fact that the Sunday of the clock changes had more or fewer hours than other days. The shortening of the Sunday of the Spring clock change was adjusted for by dividing the number of events by 23 and multiplying by 24, whilst the lengthening of the Sunday of the Autumn clock change was accounted for by dividing the number of events by 25 and multiplying by 24.

Analyses were performed in Stata version 18. Data cleaning and analysis code written by MdeL was reviewed by K.B. The complete code is available at https://github.com/MeldeLange/dst_cprd. This study meets all five CODE-EHR framework standards for using structured healthcare data in clinical research^30^ (see supplementary text S5 for the completed checklist). It was written according to the STROBE-RECORD checklist for observational studies using routinely collected health data^31^ (see supplementary text S6).

#### Secondary analyses

As a secondary analysis we ran the primary analysis stratified by age, sex, IMD and incident versus prevalent cases. Anxiety, depression, psychiatric conditions in A&E, self-harm, sleep disorders and road traffic injuries were also stratified by alcohol status. Eating disorders was also stratified by BMI. We also stratified analyses for acute CVD by BMI, alcohol status, smoking status, systolic and diastolic blood pressure, and CVD subgroup (see supplementary table S2 for details of how these covariates were derived). We used Cochran’s Q to test for differences between strata.

We also examined the number of health events over different post-clock change periods (see supplementary figures S1b-e). Specifically, we compared: 1) Health events on the individual days of the Sunday to Saturday in the week after the clock change to a control period consisting of the same day in the four weeks before the clock change and weeks 2-4 after the change. This aimed to examine whether the clock changes were changing the daily timing of events rather than the total number of events across the whole week. 2) Health events on the Monday to Friday of the first week after the clock change to a control period of the Monday to Friday in the four weeks before the clock change and weeks 2-4 after the change. 3) Health events in the first two weeks after the clock changes to a control period of the four weeks before the change and weeks three to four afterward. 4) Health events in the four weeks after the clock change to a control period of the four weeks before the clock changes.

#### Sensitivity analyses

We conducted two sensitivity analyses to examine the effect of an individual having more than one event for the same outcome in the eight weeks surrounding one particular clock change. 1) We excluded the events of anyone with an extreme number of events (≥20) in a particular eight-week period from the sub-dataset for those eight weeks. We only kept one randomly selected event for each remaining person in those eight weeks. 2) For each clock change, we excluded anyone with an event for that outcome in weeks 12 to five before the clock change (i.e. the eight weeks before our eight-week study period). We then kept just the first event in the eight-week study period for each individual.

#### Negative control

We also performed a negative exposure analysis to test whether our results were specific to the clock changes, or were due to seasonality more generally. Here we repeated the primary analysis, but defined the exposed week as the fourth week before the clock change. The control period consisted of weeks 8 to 5 and weeks 3 to 1 before the clock change (see supplementary figure S1f).

#### Patient and public involvement

In this study, we analysed routinely collected electronic health records data. Patients and the public were not directly involved in the design, conduct, reporting, or dissemination plans. However, CPRD provides lay summaries of all approved protocols and details of all published studies on its website (www.cprd.com). We will disseminate study results to the public via social media and press releases.

## RESULTS

### Participant characteristics

In our study, 57% of participants were female, 64% were aged 40 years or over, and 26% were in the top quintile of deprivation (see Table 1). As our study only includes people with a health event, our study population is not comparable to the general population. However, the fact that the characteristics of individuals experiencing an event in Spring and Autumn are similar suggests that our assumption that the characteristics of people experiencing events does not change over time, holds.

**Table 1.** Sample characteristics at each patient’s start of follow-up*.

|  | **Total Sample\*\*** | **Spring clock change** | **Autumn clock change** |
| --- | --- | --- | --- |
| **Total (n)** | 683,809 | 410,763 | 424,264 |
| **Sex** |  |  |  |
| Male | 294,482 (43.1%) | 174,981 (42.6%) | 180,635 (42.6%) |
| Female | 389,318 (56.9%) | 235,775 (57.4%) | 243,622 (57.4%) |
| Missing | 9 (0.0%) | 7 (0.0%) | 7 (0.0%) |
| **Age Category** |  |  |  |
| <10 | 14,323 (2.1%) | 7,148 (1.7%) | 8,260 (1.9%) |
| 10-19 | 51,783 (7.6%) | 28,783 (7.0%) | 31,263 (7.4%) |
| 20-29 | 86,196 (12.6%) | 49,808 (12.1%) | 52,911 (12.5%) |
| 30-39 | 95,629 (14.0%) | 56,299 (13.7%) | 59,456 (14.0%) |
| 40-49 | 105,432 (15.4%) | 63,531 (15.5%) | 65,852 (15.5%) |
| 50-59 | 88,482 (12.9%) | 53,497 (13.0%) | 54,900 (12.9%) |
| 60-69 | 88,393 (12.9%) | 54,169 (13.2%) | 55,425 (13.1%) |
| 70-79 | 88,569 (13.0%) | 56,383 (13.7%) | 56,369 (13.3%) |
| 80-89 | 56,143 (8.2%) | 35,696 (8.7%) | 34,671 (8.2%) |
| >=90 | 8,859 (1.3%) | 5,449 (1.3%) | 5,157 (1.2%) |
| **Index of Multiple Deprivation** |  |  |  |
| Most Deprived | 175,621 (25.7%) | 107,208 (26.1%) | 110,765 (26.1%) |
| Rest | 507,650 (74.2%) | 303,198 (73.8%) | 313,167 (73.8%) |
| Missing | 538 (0.1%) | 357 (0.1%) | 332 (0.1%) |

\*Start of follow-up =latest date of a patient's start date in CPRD Gold or the study start date (2.3.2008). Index of Multiple Deprivation is from 2019.
\*\*Patients can appear in both Spring and Autumn analyses, hence total sample is less than the Spring and Autumn samples combined.

### Primary analysis

Supplementary figures S2-S17 show the total number of events per day over the 8-week clock change periods for each health outcome over the study period (years 2008-2019). There was little evidence of seasonal patterns, such as a general increase or decrease in event numbers over the eight-week period. However, the overriding pattern seen in the graphs was that the number of health events varies according to the day of the week, with the largest variation being between weekdays and weekends. This is most likely the artifact of the healthcare system, as people are less likely to visit their GP on a weekend.

In Spring, the number of major acute CVD events increased by 2% in the week after the clock changes compared to the control period (incidence rate ratio [IRR] 1.02, 95% CI 1.005 to 1.030) (see figure 2 and supplementary table S4). However, both sensitivity analyses conducted to explore the effect of individual patients having multiple events in the same eight-week clock change period indicated that this result may have been driven by some individuals having multiple events and is, therefore, not a true effect of the clock change (see supplementary tables S5 and S6). No differences in event numbers were seen in the primary analysis for the other health outcomes in the full week after the Spring clock change compared to the control period. The results of the sensitivity analysis for these outcomes was very similar to the primary analysis, suggesting that people having multiple events did not affect these results.

**Figure 2.**
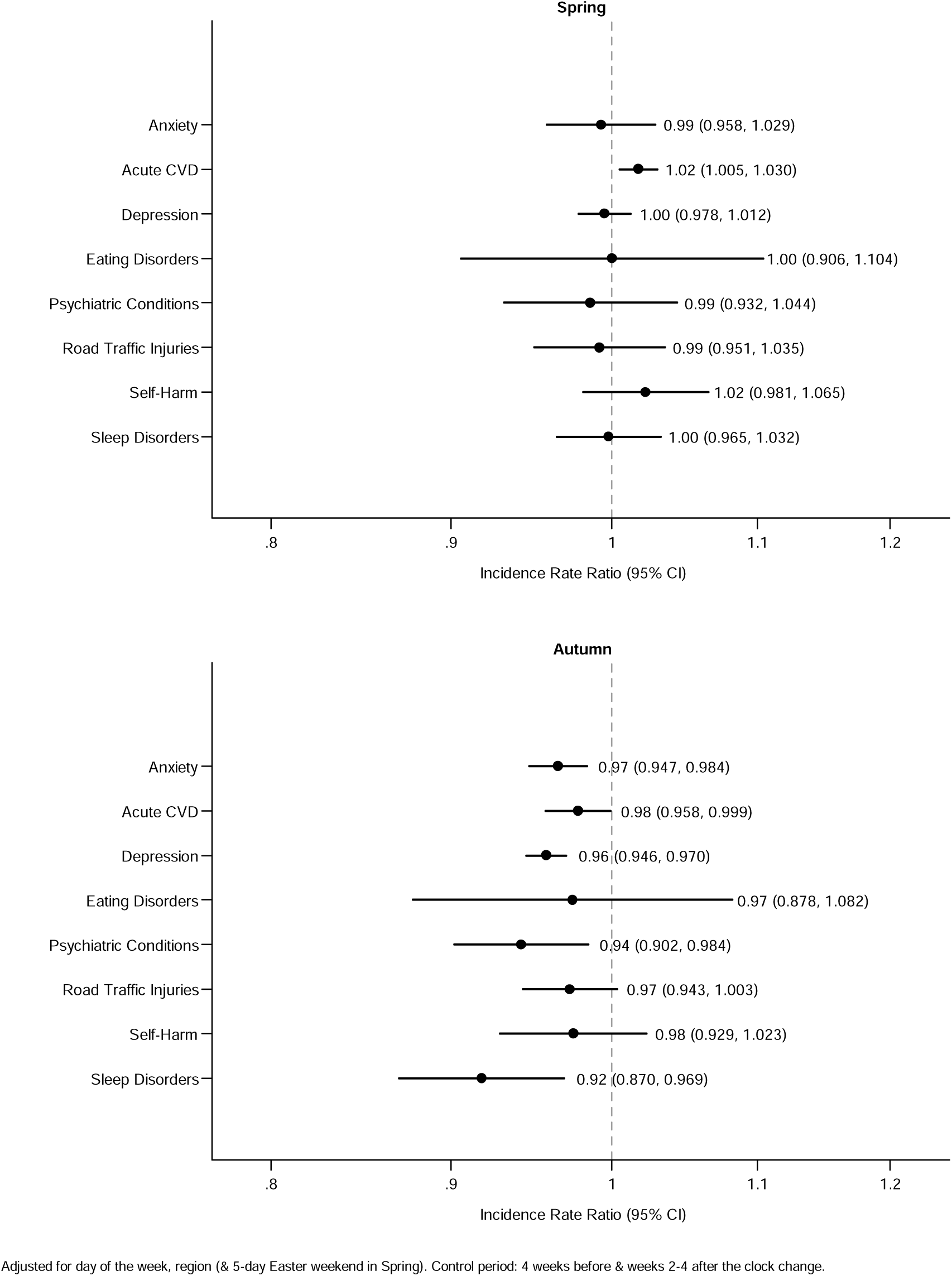
Adjusted rate ratios of the number of health events in the week after the Spring and Autumn clock changes compared to the control period (England, 2008-2019)

In the week after the Autumn clock change, there was a reduction in the number of events for anxiety disorders (-3%) (IRR 0.97, 95% CI 0.95 to 0.98), major acute CVD (-2%) (IRR 0.98, 95% CI 0.96 to 0.999), depression (- 4%) (IRR 0.96, 95% CI 0.95 to 0.97), psychiatric conditions in A&E (- 6%) (IRR: 0.94, 95% CI 0.90 to 0.98) and sleep disorders (-8%) (IRR 0.92, 95% CI 0.87 to 0.97) (see figure 2 and supplementary table S4).

### Secondary analyses

The decline in event rates for anxiety, depression and psychiatric conditions in A&E in the week after the Autumn clock change was driven by those in the youngest age categories (see supplementary table S7 for stratified results and Cochran’s Q test of heterogeneity p values). For example, the number of psychiatric conditions events recorded in A&E fell by 10% in those aged 10-35 years (IRR: 0.90, 95% CI 0.843 to 0.954), whilst there was no difference in those aged over 35 (IRR: 0.99, 95% CI 0.930 to 1.044). In addition, differences by sex and alcohol consumption were seen for psychiatric conditions in A&E. Women experienced an 11% decrease in psychiatric conditions in the week after the Autumn clock change (IRR: 0.89, 95% CI 0.835 to 0.947), whilst men exhibited no difference to the control period. In addition, current drinkers and those with missing alcohol status experienced a 5% and 13% decrease in psychiatric events in the week after the Autumn clock change, respectively, whereas for non/ex-drinkers there was weak evidence against the null hypothesis.

Our secondary analysis showed considerable variability in daily event numbers over the first week after the clock change (see supplementary table S4). In many cases, increases or decreases seen on particular days did not persist for the whole week, suggesting that the clock changes could have affected the timing of events across the week, rather than the total number of events within the week. For example, the number of depression events dropped by 8% on the Sunday of the Spring clock change, whilst the number of road traffic injuries increased by 2% on the same day, However, there was no effect for either of these outcomes over the entire first week.

Overall, the results of our analysis of events on the Monday-Friday in the first week after the clock changes were highly consistent with our primary analysis. Analyses comparing the number of events in the two and four weeks after the clock changes to their respective control periods (two-week analysis control period: 4 weeks before the clock change and weeks 3-4 after; four-week analysis control period: 4 weeks before the clock change) found few differences (See supplementary table S4). However, event rates for depression were 2% lower than the control period in the two weeks after the Autumn clock change and the number of sleep disorder events recorded was 7% lower than the control period in the two weeks, and 3% lower in the four weeks, after the Autumn clock change.

### Negative control

Our negative exposure analysis (where the exposed week was the fourth week before the clock change and the control period consisted of weeks 8 to 5 and weeks 3 to 1 before the clock change) detected little evidence of the downward trends in Autumn seen in the main analysis (see supplementary table S8). In Spring it detected an increase in sleep disorders in the week after the negative exposure, which was not found in our main analysis.

## DISCUSSION

### Principal findings

We found a reduction in the number of events (diagnoses or symptoms and accompanying prescription for mental health conditions in primary care) recorded for multiple health conditions in the week after the Autumn clock change. This included anxiety, major acute CVD, depression, psychiatric conditions in A&E and sleep disorders. The reduction in sleep disorders persisted for four weeks, although the effect attenuated over that period. We found little evidence that the Spring clock changes affected the number of health events recorded in the week after the change.

### Comparison with other studies

The latest meta-analysis of the effects of the clock changes on AMI reported a 4% increase after the Spring clock change, but there was substantial heterogeneity in study results and designs^3^. We found initial evidence of a 2% increased risk of acute CVD in the week after the Spring clock change which was not supported by sensitivity analyses examining the effect of individuals having multiple events. Our results may differ because we included primary care data, used data from only England, with its unique geographical location and healthcare system, and included other CVD conditions, such as stroke. Studies have pointed to the clock changes affecting the day of the week strokes occur rather than the total number of events across the post-clock change week^7^. In line with our results, other studies have found no difference in overall CVD hospital admissions or deaths after the Spring change.^32, 33^

Our null findings for road traffic injuries in Spring align with a systematic review which reported inconsistent findings^9^. Our results are also consistent with the only previous study to include data from England which detected a very small absolute reduction in police-recorded road traffic casualties after the Spring clock change (-0.003%)^10^. Meanwhile, our results for depression are supported by studies that have also found little evidence of an effect of the Spring clock change on depression and other mental health conditions.^5, 12, 28^

A Danish study found that the Autumn change was associated with an 11% increase in unipolar depressive episodes in hospitals^5^. This discrepancy with our results (4% decrease) may be because we included primary care data in our analyses, which captures less acute events. There could also be some misclassification in our study due to there being a delay between people experiencing depression and seeing a clinician in primary care. In addition, the contrasting results may be due to disparities in sunrise and sunset times between England and Denmark as a result of their geographical location.

The meta-analysis of AMI mentioned above reported that the Autumn clock change had no effect.^3^ Other studies, however, have reported a 30% reduction in cardiac arrests^34^, a 7.5% decrease in hospital admissions due to CVD^6^ and a 3% reduction in all-cause mortality after the Autumn change.^35^ Our 2% decrease in CVD events after the Autumn clock change is therefore generally in line with estimates reported by the existing literature. The effect of the clock changes on sleep disorders and anxiety have not been explored previously, therefore our evidence of a decline in these conditions after the Autumn clock change represents a novel contribution to the literature.

### Strengths and weaknesses of this study

This comprehensive examination of the effects of the DST clock changes on the health of the English population benefitted from access to 12 years of data from a large dataset of linked electronic health records, that is broadly representative of the UK population.^22^ Our inclusion of primary care, hospital admission and A&E data meant that our study captured a more complete picture of the effect of the clock changes on health services demand than previous studies. Furthermore, the breadth of data available meant that we were able to examine multiple different health outcomes (some not examined in this context before) in the same dataset. In addition, we conducted several sensitivity analyses to evaluate the robustness of our results.

The quality of routinely collected health data relies on the accuracy and completeness of data recorded by health professionals.^36^ CPRD Gold data have been found to have high levels of validity,^36^ and we only used data from GP practices considered to be ‘up to standard’ by CPRD. Nonetheless, mental health outcomes and sleep disorders are particularly difficult to define in electronic health records, with wide variability in the definitions and codes used by researchers to identify cases.^37^ For this reason, we used definitions and code lists for these outcomes created by an earlier study after the authors systematically reviewed previously used code lists.^38^ We have also fully reported the outcome definitions and code lists we used in this study (See Supplementary Text S3 and https://github.com/MeldeLange/dst_cprd/code_lists respectively).

However, electronic health records only include health events where the individual seeks medical help. It is possible that more subtle effects on health, such as a slight dip in mood, were not captured in this study. Additionally, there could be potential for misclassification in primary care data whereby people diagnosed after the clock change may have been experiencing symptoms for non-urgent conditions before the change but only sought or received help after the change.

Whilst we controlled for day of the week, geographical region, and the Easter weekend in our analyses, it was not possible to adjust for the competing exposure of school half-term holidays, as these vary widely throughout England. It is also possible that our stratified analyses lacked statistical power, particularly for less common health outcomes such as eating disorders.

### Implications for policy

Our study contributes to the ongoing debate over England’s clock change policy. It provides evidence that, in England, the Autumn clock change reduces demand for NHS services for CVD, sleep disorders and mental health disorders. The reductions in health events reported in this study are relatively small in terms of percentages (decreases of 2-8%). However, because the clock changes affect the whole population of England, the underlying change in number of events is high. Even small effects can have substantial implications for demand for NHS appointments and prescriptions. Furthermore, reductions in conditions such as sleep disorders could prevent people going on to develop other mental and physical health problems such as diabetes^39^, depression^40^, dementia^41^ and cardiovascular disease^42^.

The clock changes are believed to influence health via a combination of sleep and light. Examining the mediators through which the clock changes affect health was not possible in our study as we did not have data on sleep or light exposure. However, the extra sleep gained over the Autumn clock change^25, 43, 44^ could reduce CVD risk by helping to control cortisol levels, inflammation, blood pressure and insulin sensitivity.^45^ Sufficient sleep is also vital for good mental health.^40, 46^

The fact that we found little evidence that the Spring clock change (when people lose an hour of sleep) had an adverse effect on our health outcomes, combined with the fact that people only gain around half an hour of sleep over the Autumn clock change^25^, suggests that morning light may be particularly important. Despite the clocks moving forward, the lack of morning light immediately after the Spring clock change is theoretical, as it is still light when most people wake up.^20^ However, after the Autumn clock change people are exposed to more sunlight in the mornings and less sunlight in the evenings.^5^ This is important for resetting our circadian rhythm each day (it naturally runs just over 24 hours)^47^, improving sleep.^20^ Morning light also improves mental health, as evidenced by the fact that morning light therapy is an effective treatment for depression.^48, 49^ This could be a potential reason for the discrepancy in findings between our study and the study of depression in Denmark, as sunrise is generally earlier in England than in Denmark. As a result, people in England will benefit from a greater increase in morning sunlight after the Autumn clock change than people in Denmark.

The results of this study should be interpreted within the context of the complete literature on DST clock changes and health.^50^ Overall, the current literature suggests that the Spring clock change has a detrimental effect on health^51^, whilst we found that the Autumn clock change had some beneficial effects.

It is difficult to determine the longer-term effect of being on DST for 7 months of the year due to the competing exposure of seasonal effects.^20^ However, studies show that later sunrises and sunsets (experienced during DST) are associated with poorer sleep.^52, 53^ This is because evening light increases alertness and suppresses melatonin release, making it harder for people to fall asleep.^54, 55^ Meanwhile, evidence from Russia’s experience of permanent DST suggests that this harmed adolescents’ sleep and mood.^56^ Ultimately, permanent DST does not create more hours of daylight, it simply prioritises evening light over morning light. In contrast, permanent standard time (GMT), as recommended by the British Sleep Society^20^ and other sleep societies^57–61^, prioritises morning light and sleep. It may, therefore, be the best option for the health of the English population.

Further research should investigate the mechanisms underlying the possible reduction in mental health conditions observed after the Autumn clock change in this study. Future studies could also utilise data from countries such as Australia, where DST is only observed in certain states, to compare rates of health events under DST and non-DST conditions in geographically similar locations during the same time period. Additional work should also analyse the emerging data from countries that have recently abolished DST clock changes to monitor the long-term effects of permanent DST and standard time on health. It would also be valuable to conduct cross-country comparisons to examine whether the effect of the clock changes varies according to the timing of sunrise and sunset.

### Conclusions

In summary, the Autumn clock change reduces rates of CVD, sleep disorders and mental health disorders. As the underlying mechanisms linking the clock changes to health are sleep and light, this supports England adopting permanent GMT.

## Contributors

N.M.D and M.A.dL developed the study concept. M.A.dL performed the data analysis and drafted the first version of the manuscript. S.V.E checked the medical code, ICD10 and HES A&E disease code lists used in the analysis. K.B. conducted a review of the code use to clean and analyse the data. M.A.dL, N.M.D, R.C.R, K.T, K.B, C.Y.S and S.V.E. interpreted the data, reviewed and revised the manuscript, and approved the final version as submitted. The corresponding author attests that all listed authors meet authorship criteria and that no others meeting the criteria have been omitted.

## Ethical approval

The protocol for this study was approved by the Clinical Practice Research Datalink Research Data Governance committee (protocol No 23_002468). Individual participant consent was not required.

## Data sharing

This study is based in part on data from the Clinical Practice Research Datalink obtained under licence from the UK Medicines and Healthcare products Regulatory Agency. The data is provided by patients and collected by the NHS as part of their care and support. The ONS is the provider of the ONS Data contained within the Dataset. HES and ONS data, copyright © (2021/2022), re-used with the permission of The Health & Social Care Information Centre. All rights reserved. The interpretation and conclusions contained in this study are those of the authors alone. The raw data used in this study is not publicly available and cannot be shared. Access to CPRD and linked data is subject to protocol approval via CPRD’s Research Data Governance (RDG) Process. See https://www.cprd.com/data-access for further details. Full disease code lists and Stata code for analyses are available at https://github.com/MeldeLange/dst_cprd

## Transparency

The corresponding author (MdeL) affirms that the manuscript is an honest, accurate, and transparent account of the study being reported; that no important aspects of the study have been omitted; and that any discrepancies from the study as originally planned (and, if relevant, registered) have been explained.

## Dissemination to participants and related patient and public communities

The findings from this study will be shared with academics and the public through presentations at sleep and epidemiology conferences, as well as via University of Bristol press releases and social media coverage at the time of publication.

