## supplementary for "The Effects of Daylight Saving Time Clock Changes on Mental and Physical Health in England: Evidence from the Clinical Practice Research Datalink (CPRD)"

**Supplementary material**

### ****Text S1: CPRD Protocol 22/002468****

CPRD Research Data Governance (RDG) Application Template

**ALL APPLICATIONS MUST BE COMPLETED AND SUBMITTED VIA THE CPRD ELECTRONIC RESEARCH APPLICATION PORTAL (eRAP)** [**www.erap.cprd.com**](http://www.erap.cprd.com)

**Applicants may use this template offline to prepare their research application, prior to submission on eRAP. Applicants must also read CPRD’s Research Data Governance (RDG) Guidance on how to complete their application found on the eRAP landing page under Related resources (** [**https://www.erap.cprd.com/**](https://www.erap.cprd.com/) **)**

**GENERAL INFORMATION ABOUT THE PROPOSED RESEARCH STUDY**

**Study Title (Max. 255 characters including spaces)**

**Estimating the Effects of Daylight Saving Time Clock Changes on Cardiovascular Outcomes, Depressive Symptoms and Road Traffic Injuries in England**

**Research Area** (place ‘**X**’ in all boxes that apply)

| Drug Safety |  | Economics |
| --- | --- | --- |
| Drug Utilisation |  | Pharmacoeconomics |
| Drug Effectiveness |  | Pharmacoepidemiology |
| Disease Epidemiology | x | Methodological |
| Health Services Delivery |  |  |

**Does this protocol describe an observational study using purely CPRD data?**

| Yes | **x** | No |
| --- | --- | --- |

**Does this protocol involve requesting any additional information from GPs, or contact with patients?**

| Yes |  | No | **x** |
| --- | --- | --- | --- |

If yes, provide the reference number:

**Chief Investigator**

| Title: | Professor | Mrs |
| --- | --- | --- |
| Full name: | Kate Tilling | Melanie de Lange |
| Job title: | Professor of Medical Statistics and MRC Investigator | PhD Student |
| Affiliation/organisation: | University of Bristol | University of Bristol |
| Email address: | | |
| CV Number (if applicable): | n/a | n/a |
| Will this person be analysing the data? (Y/N) | N | Y |

**Corresponding Applicant**

| Title: | Mrs |
| --- | --- |
| Full name: | Melanie de Lange |
| Job title: | PhD Student |
| Affiliation/organisation: | University of Bristol |
| Email address: | |
| CV Number (if applicable): | n/a |
| Will this person be analysing the data? (Y/N) | Y |

**List of all investigators/collaborators**

| Title: | Dr |
| --- | --- |
| Full name: | Kate Birnie |
| Job title: | Senior Research Fellow |
| Affiliation/organisation: | University of Bristol |
| Email address: | |
| CV Number (if applicable): | 465_20 |
| Will this person be analysing the data? (Y/N) | Y |

| Title: | Professor |
| --- | --- |
| Full name: | Neil Davies |
| Job title: | Professor of Medical Statistics |
| Affiliation/organisation: | University College London |
| Email address: | |
| CV Number (if applicable): | 456_15CEL |
| Will this person be analysing the data? (Y/N) | N |

| Title: | Dr |
| --- | --- |
| Full name: | Sophie Eastwood |
| Job title: | Clinical Research Fellow |
| Affiliation/organisation: | University College London |
| Email address: | |
| CV Number (if applicable): | n/a |
| Will this person be analysing the data? (Y/N) | N |

**ACCESS TO THE DATA**

**Sponsor of the study**

| Institution/Organisation: | Medical School, University of Bristol |
| --- | --- |
| Address: | Oakfield House, Oakfield Grove, Clifton, Bristol, BS8 2BN |

**Funding source for the study**

| Same as Sponsor? | Yes |  | No | X |
| --- | --- | --- | --- | --- |
| Institution/Organisation: | Wellcome Trust | | | |
| Address: | Gibbs Building, 215 Euston Road, London, NW1 2BE | | | |

**Institution conducting the research**

| Same as Sponsor? | Yes | X | No |
| --- | --- | --- | --- |
| Institution/Organisation: |  | | |
| Address: |  | | |

**Data Access Arrangements**

Indicate with an ‘**X**’ the method that will be used to access the data for this study:

| Study-specific Dataset Agreement | X |
| --- | --- |

| Institutional Multi-study Licence |
| --- |
| Institution Name |
| Institution Address |

Will the dataset be extracted by CPRD?

| Yes | X | No |
| --- | --- | --- |

If yes, provide the reference number:

**Data Processor(s):**

| Processing | X |  |
| --- | --- | --- |
| Accessing | X |  |
| Storing | X |  |
| Processing area (UK/EEA/Worldwide) | | UK |
| Organisation name | | Medical School, University of Bristol |
| Organisation address | | Oakfield House, Oakfield Grove, Clifton, Bristol, BS8 2BN |

**INFORMATION ON DATA**

**Primary care data** (place ‘**X**’ in all boxes that apply)

| CPRD GOLD | X | CPRD Aurum |
| --- | --- | --- |

**X**

Reference number (if applicable):

**Please select any linked data or data products being requested**

**Patient Level Data** (place ‘**X**’ in all boxes that apply)

| ONS Death Registration Data |  | NCRAS Cancer Registration Data |
| --- | --- | --- |
| HES Admitted Patient Care | X | NCRAS Systemic Anti-Cancer Treatment (SACT) data |
| HES Outpatient |  | NCRAS National Radiotherapy Dataset (RTDS) data |
| HES Accident and Emergency | X | Second Generation Surveillance System (SGSS, COVID-19) |
| HES Diagnostic Imaging Dataset |  | COVID-19 Hospitalisations in England Surveillance System (CHESS) |
| Mental Health Data Set (MHDS) |  |  |
| CPRD Mother Baby Link |  |  |
| Pregnancy Register |  |  |

**Area Level Data** (place ‘**X**’ in one Practice / Patient level box that may apply)

| **Practice level (UK)** |  | **Patient level (England only)** |  |
| --- | --- | --- | --- |
| Practice Level Index of Multiple Deprivation |  | Patient Level Index of Multiple Deprivation | X |
| Practice Level Index of Multiple Deprivation Domains |  | Patient Level Index of Multiple Deprivation Domains |  |
| Practice Level Carstairs Index for 2011 Census (Excluding Northern Ireland) |  | Patient Level Carstairs Index for 2011 Census |  |
| 2011 Rural-Urban Classification at LSOA level |  | 2011 Rural-Urban Classification at LSOA level |  |
|  |  | Patient Level Townsend Score |  |

Reference / Protocol number (where applicable):

**Are you requesting linkage to a dataset not listed above?**

| Yes |  | No | **X** |
| --- | --- | --- | --- |

If yes, provide the Non-Standard Linkage reference number:

**Does any person named in this application already have access to any of these data in a patient identifiable form, or associated with an identifiable patient index?**

| Yes |  | No | **X** |
| --- | --- | --- | --- |

If yes, provide further details:

**PART 2: PROTOCOL INFORMATION**

**Applicants must complete all sections**

**Study Title**

**Estimating the Effects of Daylight Saving Time Clock Changes on Cardiovascular Outcomes, Depressive Symptoms and Road Traffic Injuries in England**

**Lay Summary (Max. 250 words)**

**Exploring the effects of Daylight Saving Time (DST) Clock Changes on Cardiovascular Disease, Depression and Road Traffic Injuries in England**

There is growing evidence that daylight saving clock changes (the clocks moving one hour forward in spring and one hour back in autumn) may have adverse effects on people’s health due to sleep deprivation and the disruption of biological rhythms. For example, studies outside of England have shown that the number of depressive episodes, heart attacks, strokes and fatal road traffic accidents increases in the weeks after the clock changes. This has prompted countries such as the US and the EU to vote to end DST clock changes. However, it is unclear whether DST will be abolished in England.

The purpose of this research is to estimate the effect of daylight savings time clock changes on the health of the English population. To do this we will compare the number of GP and hospital visits for cardiovascular disease, depression and road traffic injuries in the 4 weeks directly before and after the clock changes.

This data will provide evidence as to whether England should abolish DST clock changes. It will also offer insights into the wider effects of sleep and circadian disruption on mental and physical health.

**Technical Summary (Max. 300 words)**

Growing evidence suggests that daylight saving time (DST) clock changes (one hour forward in spring and one hour back in autumn) may have adverse effects on population health, likely via sleep deprivation and circadian disruption. For example, several studies using small datasets from outside England have reported increased incidence of depressive episodes, myocardial infarction, atrial fibrillation, strokes and fatal traffic accidents in the weeks immediately following the clock changes. This has prompted the US and the EU to vote to end DST clock changes. However, it is currently unclear whether England will do the same.

The aim of this study is to estimate the effects of DST transitions on cardiovascular disease, depression and road traffic injuries in the English population. Using CPRD and HES data for 2008-2022, regression discontinuity analysis will be used to compare the average number of primary and secondary care visits for these health outcomes in the 4 weeks before and after the spring and autumn clock changes. Regression discontinuity designs are a statistical method that uses a specific threshold or cut-off point (here the time of DST clock change) to estimate the causal, real-world, effects of policies. Differences in the number of visits will be compared in the 1- (modelled daily), 2- and 4-week periods before and after the clock changes.

Research findings will help to quantify the effects of DST clock changes on English population health by estimating the number of incident cases of disease and recurrent events attributable to DST clock changes. The results will be triangulated to formulate policy recommendations as to whether the English government should abolish DST clock changes. More broadly, the research will also offer insights into the wider effects of sleep and circadian disruption on mental and physical health.

**Outcomes to be Measured**

**Primary outcomes**

All primary/secondary care visits (incident & prevalent cases) in the 1- (modelled daily), 2- and 4-week periods before and after the clock changes, stratified by age (10 year age bands), for:

Depression (aged ≥10*).

Road traffic injuries (all ages).

Cardiovascular disease (aged ≥40**).

*Age ≥10 was chosen to ensure we capture adolescents who may be disproportionately affected by the sleep deprivation caused by the clock changes due to a shift towards having a later chronotype (going to bed and getting up later) during adolescence.

**The age at which people become eligible for their NHS health check and are considered at higher risk of cardiovascular disease.

**Secondary outcomes**

All primary/secondary care visits (incident & prevalent cases) in the 1- (modelled daily), 2- and 4-week periods before and after the clock changes, aged ≥10, stratified by age (10 year age bands), for:

Sleep disorders. This analysis will be exploratory, and will only be conducted if it is possible to adequately define sleep disorders within CPRD data.

Other mental health (anxiety & self-harm) & eating disorders.

The number of subsequent hospitalisations/referrals for depression/cardiovascular disease, or deaths amongst those visiting primary/secondary care for depression/cardiovascular disease in the 1-, 2- and 4-week periods before and after the clock changes.

**Detailed definition of outcomes**

Cardiovascular disease, road traffic injuries, self-harm and eating disorders will be defined using ICD10, Read codes and HES A&E codes.

Depression and anxiety will be defined using ICD-10, Read codes and HES A&E codes.

Read codes alone will be used to define depression/anxiety if the code is specific enough (see code lists C1 and C6 for depression/anxiety diagnoses).

However, where Read codes relate to symptoms of depression/anxiety (see code lists C2 and C7 for depression/anxiety symptoms) that are not considered adequate to define depression/anxiety, patients will only be counted as having depression/anxiety if they have been prescribed a drug used to treat depression within 90 days.

Sleep disorders will be defined using ICD-10 and Read codes.

Sleep disorders are difficult to define which means that most Read codes are not specific and relate to symptoms rather than diagnoses. Patients will therefore only counted as having a sleep disorder if they have a Read code for a sleep disorder diagnosis or symptom and have been prescribed a drug used to treat sleep disorders within 90 days.

See Appendix A for dates of clock changes, Appendix B for preliminary ICD10 codes, Appendix C for preliminary Read v2 codes, Appendix D for preliminary prescription codes and Appendix E for HES A&E codes.

*Notes to reviewer:

Having death as an outcome would be interesting, but unfortunately we do not have the budget to buy the linked ONS data.

Looking at the different severities of presentation would also be interesting but would be challenging to assess and other studies have struggled to do this using CPRD data.

**Objectives, Specific Aims and Rationale**

**Aims and objectives**

The overall aim of this research is to estimate the effect of daylight savings time clock changes on the health of the English population.

More specifically we want to:

Estimate the effects of DST clock changes on depression, road traffic injuries and cardiovascular disease.

Gain insights into the broader effects of sleep deprivation and circadian disruption on mental and physical health, including on sleep disorders, anxiety, self-harm, eating disorders.

Make policy recommendations as to whether England should abolish DST clock changes.

**Rationale**

Knowledge/information to be gained: By quantifying the health effects of DST clock changes on the English population the knowledge gained from the study will help to build an evidence base which can then be used to inform public health policy relating to whether the England should abolish DST clock changes. The findings will also improve our understanding of the effects of sleep deprivation and circadian disruption on mental and physical health.

Primary hypothesis to be tested: Primary/secondary care visits for depression, road traffic injuries and cardiovascular disease will be higher in the weeks after DST clock changes than in the weeks prior to the clock changes.

How achievement of objectives will further the research aim: In achieving our objectives we will have conducted one of the largest studies of the effects of DST on health in the UK to date. As well as informing English policy, the findings also have relevance to other countries considering abolishing DST clock changes. More widely, the research will improve our understanding of how sleep and circadian rhythms affect physical and mental health.

**Study Background**

Switching to daylight saving time (DST) involves moving the clocks one hour forwards in spring and one hour backwards in autumn^1^. It was first introduced during World War One^2^ as a way of maximising exposure to daylight during the working day^1^ and subsequently reducing energy use^3^. DST is now in operation in over 70 countries, including the UK, and affects over a quarter of the world’s population^1^,^4^.

There is growing evidence that DST clock changes may have adverse effects on population health^1^, likely via effects of sleep deprivation and circadian disruption^5^. For example, several studies using small datasets from outside of England have reported increased incidence of depressive episodes^1^, myocardial infarction (MI)^6^, atrial fibrillation^7^, strokes^8^ and fatal traffic accidents^9^ in the weeks immediately following the clock changes. The likely adverse health effects of DST transitions has prompted calls for DST to be abolished and a number of countries are currently reconsidering its use^3,10^. Most prominently, in March 2019 the European Union voted to end DST after 2021^11^. However, it is currently unclear whether England will do the same.

Although previous studies have generally found consistent effects of DST on health outcomes, most have used relatively small datasets limiting the precision of effect estimates. For example, a 2018 review noted that the combined number of observations from 6 previous studies of DST and MI amounted to less than 90,000 individuals^12^. Furthermore, it is not clear to what extent previous findings can be generalised across countries with different climates, lifestyles and healthcare systems^6^.

**Study Type**

This study will be descriptive in that it will describe and compare the frequency of primary and secondary care visits for depression, road traffic injuries, cardiovascular disease, sleep disorders, anxiety, self-harm and eating disorders within certain time periods.

It will also be hypothesis testing in that we will be looking at whether there is a discontinuity in, and statistical difference between, the number of visits before and after the clock changes. Our hypothesis is that there will be a difference and that the number of visits will be higher in the weeks after the clock changes.

**Study Design**

This research utilises a regression discontinuity design. Regression discontinuity designs are a statistical method that uses a specific threshold or cut-off point to estimate the causal effects of interventions. They are particularly valuable in evaluating the real world effects of policies, which inherently cannot be examined in a randomised control trial^13^. Here the DST clock changes act as a cut-off point determining whether people are exposed (the weeks after the clock changes) or unexposed (weeks before the clock changes). A recent meta-analysis comparing the results of regression discontinuity studies and randomised control trials concluded that regression discontinuity studies have high levels of interval validity^14^.

We will also conduct stratified subgroup analysis to explore whether the effects of the clock changes are modified by factors such as year, incident vs. prevalent case, sex, age, ethnicity, socioeconomic position, alcohol, smoking, BMI, blood pressure, co-morbidities and disease subgroups.

**Feasibility counts**

Table 1 presents feasibility counts for our 3 primary outcomes. Counts for depression and cardiovascular disease were taken from existing CPRD publications. Feasibility counts for road traffic injuries are based on the November 2022 CPRD Gold primary care database.

Table 1: Estimated Sample Sizes of Patients in England Eligible for Linkage to HES datasets

| **Outcome** | **Data Source** | **Average cases per year** | **Average cases per week per year** | **Average cases in 8 week period per year*** | **Cases in 8 week period for our 15 year study** |
| --- | --- | --- | --- | --- | --- |
| Road traffic injuries (all cases) | CPRD Gold** | 5,275 | 101 | 812 | 12,174 |
| Cardiovascular disease*** (incident only) | Gho et al (2018), Wu et al. (2022), Yang et al. (2021), Lawson et al. (2021)^15-18^ | 19,489 | 375 | 2998 | 44,974 |
| Depression  (all cases) | Kendrick et al. (2015)^19^**** | 22,020 | 423 | 3388 | 50,815 |

*We are looking at the 8 week period of 4 weeks before & after each clock change.

**Identified using CPRD Gold medical code 279: Motor vehicle traffic accidents (MVTA). These estimates do not include HES and are likely to be conservative.

***Myocardial infarction, atrial fibrillation, stroke & heart failure.

****Counts are based on 75% of the counts in the original paper which used all patients in England rather than just those with HES linkage (75% of English practices have HES linkage).

**Sample size considerations**

The minimum detectable effect size for our primary outcomes at 80% power was calculated in Stata using the power twoproportions function.

Table 2: Calculations to Estimate Minimum Detectable Effect Size at 80% Power

| **Outcome** | **Total underlying population from existing literature*** | **Cases in 4 week pre-exposure period over 15 year study** | **Prevalence (unexposed)**** | **Assumed Prevalence exposed***** | **Minimum detectable effect size (percent increase between exposed and unexposed) at 80% power** |
| --- | --- | --- | --- | --- | --- |
| Road traffic injuries | 3,383,148 | 6,090 | 0.001800099 | 0.001931686 | 7.31% |
| Cardiovascular disease | 3,383,148 | 22,485 | 0.006646177 | 0.006896073 | 3.76% |
| Depression | 3,383,148 | 25,410 | 0.007510756 | 0.007776637 | 3.54% |

*From Wu et al (2022) Patients in England aged ≥16 years, contributing data 1998-2017,eligible for HES linkage.^16^

**15-year 4-weekly cases/total population

***(Unexposed prevalence * (effect size detectable/100)) + unexposed prevalence

**Planned use of linked data (if applicable):**

The following linkages will be requested:

HES Admitted Patient Care (HES-APC)

HES A&E

Index of Multiple Deprivation (IMD), patient level

Linked HES data will be important to gain full data on our outcomes, many of which will be more likely to be captured in secondary care data. Linked CPRD and HES data will be used to identify prevalent and incident cases for subgroup analyses exploring the effects of DST clock changes on prevalent and incident cases separately (where appropriate). Patient Level Index of Multiple Deprivation data will be used to identify whether socioeconomic position is a modifier of the effect of DST clock changes on our health outcomes.

A secondary use of the linked data will be to identify significant subsequent health events (hospitalisations/referrals for depression/cardiovascular disease, or deaths) amongst those visiting primary/secondary care for depression/cardiovascular disease.

**Definition of the Study population**

a) Target population: The population of England (all ages for road traffic accidents) (aged ≥10 for depression, anxiety, self-harm, eating disorders and sleep disorders) (aged ≥40 for cardiovascular disease).

b) Recruitment period & start/end of follow-up: 2008 – 2022

Recruitment starts in 2008 as all the datasets we are using are available from then onwards (first full year of HES A&E data is 2008).

c) Inclusion criteria (based on clinical data):

1. All patients in England, eligible for data linkage, registered in the CPRD GOLD database in the 4 weeks before and after each DST clock change.

2. All ages for road traffic injuries.

Aged ≥10 for depression, sleep disorders, anxiety, self-harm and eating disorders.

Aged ≥40 for cardiovascular disease.

3. Clinical record with at least 12 consecutive months of records classified as ‘acceptable’ by the CPRD from all “up to standard” practices.

In the UK the clocks go forward 1 hour at 1am on the last Sunday in March and go back 1 hour at 2am on the last Sunday in October. Consequently, the clock changes do not occur on the same date each year. For a full list of clock change dates by year see Appendix A.

Exclusion criteria:

Patients aged <10 years for depression, anxiety, eating disorders, self-harm and sleep disorders

Patients aged <40 for cardiovascular disease

For analysis of significant subsequent health events: those with data not up to standard during the follow-up period.

d) Index date: Event date (date of visit)

e) Incident case: First ever record for this disease code. Prevalent case: Has record for this disease code prior to event date.

f) Sampling from base population: N/A.

g) Exposure windows: Exposed = The 4 weeks after the clock changes.

Unexposed = The 4 weeks before the clock changes.

h) Linked data will be subject to the same eligibility criteria.

**Selection of comparison group(s) or controls**

In this study we are comparing our exposed group (those visiting primary/secondary care after the clock change) to our unexposed group (those visiting primary/secondary care before the clock change)

Figure 1: Diagram to Show Unexposed and Exposed Periods

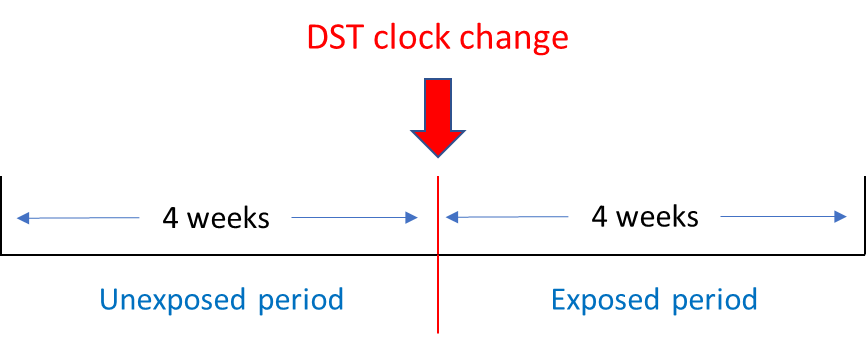

**Exposures, Outcomes and Covariates**

**Exposure periods:**

Unexposed: 4 weeks before the clock change.

Exposed: 4 weeks after the clock change.

**Primary outcomes**

All primary/secondary care (incident & prevalent cases) visits in the 1- (modelled daily), 2- and 4-week periods before and after the clock changes, stratified by age, for:

Depression (aged ≥10).

Road traffic injuries (all ages).

Cardiovascular disease (aged ≥40*.

*The age at which people become eligible for their NHS health check and are considered at higher risk of cardiovascular disease.

**Secondary outcomes**

All primary/secondary care visits (incident & prevalent) in the 1- (modelled daily), 2- and 4-week periods before and after the clock changes, aged ≥10, stratified by age, for:

Sleep disorders. This analysis will be exploratory, and will only be conducted if it is possible to adequately define sleep disorders within CPRD data.

Other mental health (anxiety & self-harm) & eating disorders.

The number of subsequent hospitalisations/referrals for depression/cardiovascular disease, or deaths amongst those visiting primary/secondary care for depression/cardiovascular disease in the 1-, 2- and 4-week periods before and after the clock changes.

**Definition of outcomes**

Cardiovascular disease, road traffic injuries, self-harm and eating disorders will be defined using ICD10, Read codes and HES A&E codes.

Depression and anxiety will be defined using ICD-10, Read codes and HES A&E codes.

Read codes alone will be used to define depression/anxiety if the code is specific enough (see code lists C1 and C6 for depression/anxiety diagnoses).

However, where Read codes relate to symptoms of depression/anxiety (see code lists C2 and C7 for depression/anxiety symptoms) that are not considered adequate to define depression/anxiety, patients will only be counted as having depression/anxiety if they have been prescribed a drug used to treat depression within 90 days.

Sleep disorders will be defined using ICD-10 and Read codes.

Sleep disorders are difficult to define which means that most Read codes are not specific and relate to symptoms rather than diagnoses. Patients will therefore only counted as having a sleep disorder if they had a Read code for a sleep disorder diagnosis or symptom and were prescribed a drug used to treat sleep disorders within 90 days.

*Note to reviewer:

Depression, anxiety and sleep disorders will not be defined based on prescription alone in order to avoid misclassification due to these drugs being prescribed for other indications.

See Appendix A for dates of clock changes, Appendix B for preliminary ICD10 codes, Appendix C for preliminary Read v2 codes, Appendix D for preliminary prescription codes and Appendix E for HES A&E codes.

All disease code lists have been reviewed by Sophie Eastwood, a primary care clinician experienced in analysing CPRD data.

Data sources:

Primary and secondary care patient and clinical records, and patient level index of multiple deprivation data.

Co-variates:

Potential confounders: the clock change coinciding with Easter Sunday, the length of day on the day of the clock change, seasonal effects and covid lockdowns (see Plans for Addressing Confounding section below).

Effect modifiers: Year, incident vs. prevalent case, sex, age, ethnicity, marital status, socioeconomic position, alcohol, smoking, BMI, blood pressure, co-morbidities and disease subgroups.

**Data/ Statistical Analysis**

We will use a regression discontinuity design to investigate whether the mean number of primary/secondary care visits for depression, cardiovascular disease, road traffic injuries, anxiety, eating disorders or self-harm (and sleep disorders if possible), differs between the 4 weeks before and after the clock changes.

Data for spring and autumn clock changes will be analysed separately, as will the data for each type of diagnosis (depression, cardiovascular disease, road traffic injuries, anxiety, eating disorders, self-harm and sleep disorders).

As well as the whole 4-week study period, differences in visits in the 1-week (modelled daily) and 2-week periods before and after the clock changes will be compared.

The same analysis plan will be used to examine differences in the number of subsequent hospitalisations/referrals for depression/cardiovascular disease, or deaths amongst those visiting primary/secondary care for depression/cardiovascular disease before and after the clock changes.

We will also conduct stratified subgroup analysis to explore whether the effects of the clock changes are modified by factors such as year, incident vs. prevalent case, sex, age, ethnicity, socioeconomic position, alcohol, smoking, BMI, blood pressure, co-morbidities and disease subgroups.

To investigate whether differences observed represent a true discontinuity rather than seasonal patterns, mean daily visits for the whole period of the 4 weeks before and after the clock changes will be plotted. Meta-regression will then be used to quantify whether the greatest discontinuity occurs at the time of the clock changes.

Sensitivity analyses excluding certain years will be conducted to address the potential confounding effects of Easter Sunday and COVID lockdowns.

Analyses will be adjusted for the fact that the Sunday following the clock changes has more or less hours than other days. The shortening of the Sunday after transition into DST will be adjusted for by multiplying the number of primary/secondary care visits by 24/23, and the lengthening of the Sunday after transition out of DST by multiplying by 24/25.

As a sensitivity analysis we will explore different methods of bandwidth selection using appropriate packages in stata^20^. We will also build in a sensitivity analysis with a different cut-off point to the clock change.

Our study is hypothesis driven and we only have 3 primary outcomes. In addition, we will focus on interpreting the effect sizes and confidence intervals rather than just p values. However, where appropriate, we will account for multiple testing when interpreting p-values, for example via False Discovery Rate adjustment or a Bonferroni correction.

*Notes to reviewer:

We are using individual level data to do our regression discontinuity analysis. Using individuals as their own controls would be tricky as once someone has had an outcome (e.g. heart attack / going to GP about depression) this makes them less likely to have the same outcome again during the study period.

Amendment:

For validation purposes, to check whether our number of events is plausible, we will calculate incident rates for our outcomes using a denominator file (stratified by age, gender & year).

Plan for addressing confounding

There are several potential confounders of the relationship between sleep and circadian rhythms before and after the clock changes, and the number of primary/secondary care visits:

Figure 2: DAG to Show Relationships between Exposure, Outcome and Confounders

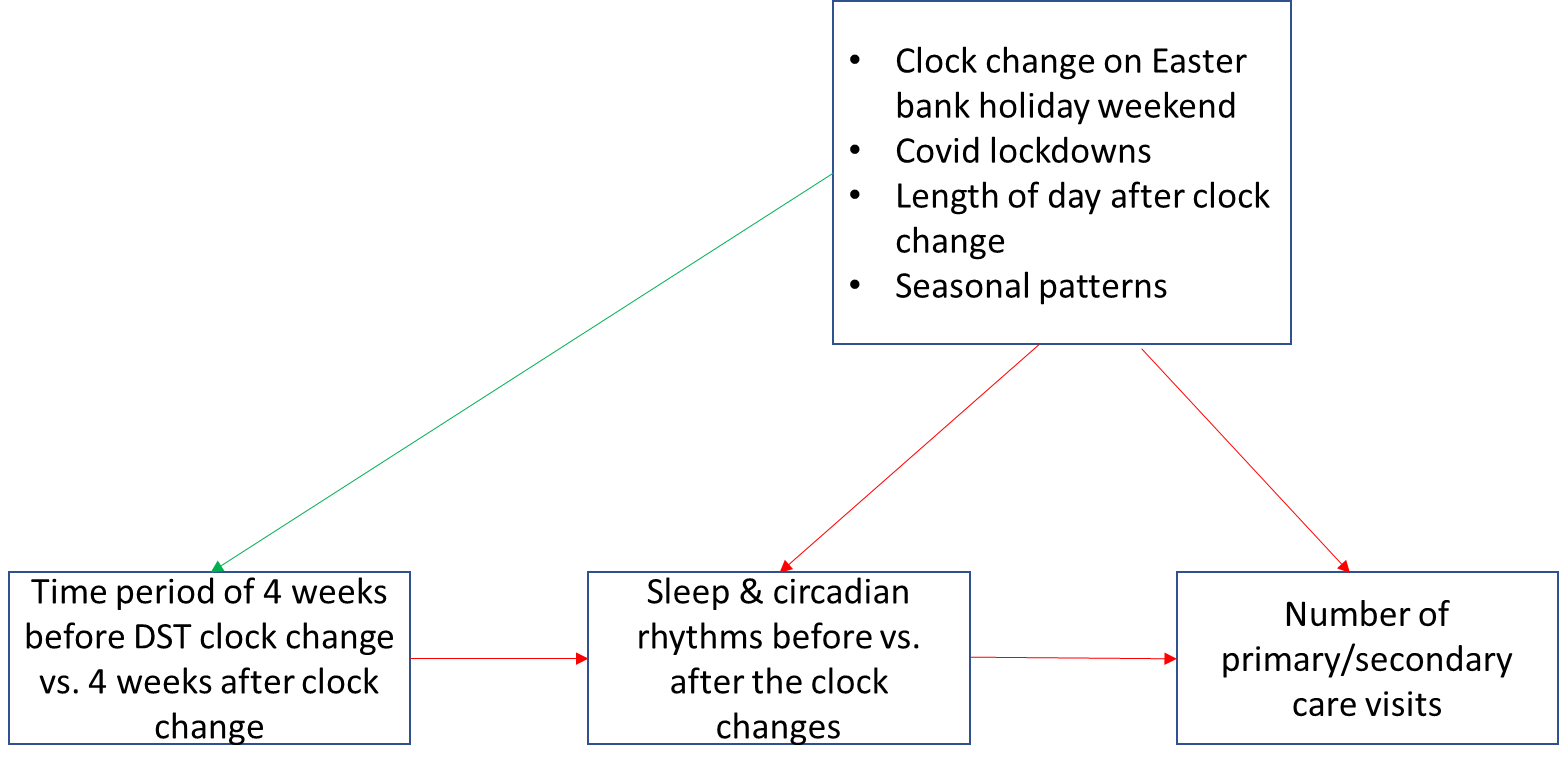

Easter – The DST clock change falls on Easter Sunday twice during our study period of 2007-2022 (See Appendix A). This could confound the acute effects of the clock change as Easter Monday is a bank holiday so people do not need to get up for work. Consequently the effects of the clock change on sleep may be delayed by a day. People are also less likely to access healthcare services on a bank holiday. To explore whether this is an issue we will conduct a sensitivity analysis excluding these years.

Length of day – The absolute number of diagnoses on the Sunday of the clock change may be confounded by the difference in the length of this day compared to all other days (-1 in spring and +1 in autumn). This will be adjusted for as described above.

Covid 19 lockdowns in 2020/21 - In 2020 the spring DST clock change was 29^th^ March. On 19^th^ March people ordered to stay at home & on 23^rd^ March lockdown measures legally came into force. The 2020 Autumn clock change was 25^th^ October. New restrictions were introduced on the 22^nd^ September and on the 5^th^ November the second national lockdown came into force in England. These lockdowns and restrictions may have affected people’s sleep (not needing to travel to work could increase sleep, stress from the situation could reduce sleep). It could also affect our health outcomes by reducing people’s access to healthcare services, as well as directly affecting rates of some outcomes such as of depression and cardiovascular disease risk (due to stress). To explore whether COVID lockdowns have affected our results we will conduct a sensitivity analysis excluding these years.

Seasonal effects - It is possible that depression increases after the autumn clock change regardless of the clock change because it is getting colder and darker (and vice versa in spring). To investigate this, mean daily visits surrounding both spring and autumn clock changes will be plotted separately to assess whether patterns show a true discontinuity or are purely seasonal trend.

Common individual level confounders (e.g. age, sex etc.) shouldn’t apply in this analysis as, although potentially associated with our outcomes, they are not associated with our exposure (sleep and circadian rhythms in the period before versus after the clock changes). They will therefore act as effect modifiers in this analysis and be examined through stratified subgroup analysis.

Plans for addressing missing data

We will investigate the extent and randomness of missing data in our exposure, outcomes and effect modifiers, and use multiple imputation to impute missing data where appropriate. Missing data is likely to be a particular issue for effect modifiers such as alcohol intake, BMI, smoking and blood pressure which are known to not be available for all patients and may not be missing at random.

Patient or user group involvement

No patients or user groups were involved in the drafting of this protocol. However, depending on our findings, we will work with Bristol’s Integrative Epidemiology Unit Public Engagement Department (www.bristol.ac.uk/integrative-epidemiology/engagement/) which engages with members of the public to share research and benefit from others' insights. The department encourages discussion about research methods and findings through a programme of activities in partnership with festivals, schools and community groups. This will enable me to engage the public, patients and policy makers in the interpretation and dissemination of our results, as well as discussing ideas for future research (see also Plans for disseminating study results section).

Plans for disseminating and communicating study results

This study will form the core of a funded 3-year PhD project. We will publish the results of the study as open-access peer-reviewed publications. Our results will be disseminated in up to six papers as detailed below:

Analysis plan for Estimating the Effects of ‘Daylight Saving Time’ Clock Changes on Health Outcomes in England.

The effect of daylight savings time transitions on depression in England.

The effect of daylight savings time transitions on cardiovascular disease in England.

The effect of daylight savings time transitions on road traffic injuries in England.

The effect of daylight savings time transitions on anxiety, eating disorders and self-harm in England.

The effect of daylight savings time transitions on sleep disorders in England (if possible).

All code will be made freely available on GitHub. We will disseminate findings through national and international conferences as are appropriate. Depending on the findings we will also explore additional options, such as media press releases and blog posts on IEUREKA (https://ieureka.blogs.bristol.ac.uk/), for wider dissemination to the public and policy makers. I will also work with PolicyBristol (www.bristol.ac.uk/policybristol) to specifically engage with policy makers and translate my research findings into clear and accessible policy briefings.

Conflict of interest statement

N/A

Limitations of the study design, data sources, and analytic methods

This study has several limitations:

This is an observational study. We will attempt to control for confounders and identify effect modifiers via, adjusting, stratified subgroup analysis and secondary analyses excluding certain years. However, any differences observed before and after the clock changes could still have arisen due to uncontrolled confounding. That said, there are unlikely to be any major individual level confounders, because DST is unlikely to be related to most individual level factors. Furthermore, we will exploit random variation in when in the year the DST change occurs to investigate this further.

GP and hospital data will only capture the most severe cases so more subtle effects of the clock changes won’t be captured in this study. However, most substantial health consequences are likely to be recorded.

There may be a delay in patients seeking and then getting medical help for non-urgent conditions such as sleep disorders and depression. This may lead to some misclassification whereby people diagnosed after the clock change were already experiencing symptoms prior to the clock change. We will evaluate this using sensitivity analyses, for example, using a fuzzy regression discontinuity design^21^.

Data (such as smoking status and BMI) are unlikely to be missing at random in healthcare data. This may make it challenging to use multiple imputation. We will run sensitivity analyses to investigate this, for example comparing the results from a complete case analysis with an analysis assuming those with missing BMI/smoking are normal weight/never smokers.

Appendix B: ICD10 Codes

Appendix B1: Depression ICD10 Codes

Appendix B2: Road Traffic Injuries ICD10 Codes

Appendix B3: Cardiovascular Disease ICD10 Codes

Appendix B4: Sleep Disorders ICD10 Codes

Appendix B5: Anxiety ICD10 Codes

Appendix B6: Self-Harm ICD10 Codes

Appendix B7: Eating Disorders ICD10 Codes

Appendix C: Read v2 codes

Appendix C1: Depression Diagnosis Read Codes

Appendix C2: Depression Symptom Read Codes

Appendix C3: Road Traffic Injuries Read Codes

Appendix C4: Cardiovascular Disease Read Codes

Appendix C5: Sleep Disorders Read Codes

Appendix C6: Anxiety Diagnosis Read Codes

Appendix C7: Anxiety Symptom Read Codes

Appendix C8: Self-Harm Read Codes

Appendix C9: Eating Disorders Read Codes

Appendix D:Prescription codes

Appendix D1: Depression Prescription Product Codes

Appendix D2: Anxiety Prescription Product Codes

Appendix D3 : Sleep Disorders Prescription Product Codes

Appendix E: HES A&E codes

Appendix F: Tables & Diagrams

Appendix F1: Feasibility Counts

Appendix F2: Sample Size Considerations

Appendix F3: Comparison Groups

Appendix F4: Confounding

Grant ID (optional)

### ****Text S2: Minor deviations from protocol****

**The linked HES A&E data provided by CPRD only contained data up to March 2020. We therefore reduced our study period from 2008-2022 to 2008-2019. This also removed the need to address the effects of covid lockdowns in our study.**

**Once we started analysing our data we discovered that there were patients who had more than one event for the same health condition within the 8-week period surrounding an individual clock change. Whilst this was not common, we decided to examine whether this was affecting our results by running the two sensitivity analyses described in the main paper.**

**We did not run stratified analyses by year, marital status, disease comorbidities or ethnicity as there were no prior indications that these would act as effect modifiers and because the sample sizes of the individual strata (and subsequently statistical power) would have been low. In addition ethnicity is poorly recorded in patients registered prior to 2006^1^.**

**We originally planned to conduct a sensitivity analysis excluding years where the clock change coincided with Easter Sunday in order to examine the potential confounding effects of Easter Sunday on the relationship between the clock changes and the number of health events. However, instead we decided to adjust all of our Spring regression analyses using an indicator variable for the five days of the Easter weekend. This maximised our sample size, allowed us to take the whole long weekend into account and considered the effects of Easter wherever it fell within our 8-week study periods around the Spring clock change.**

**In our analysis we split the data by day, year and region to account for all sources of variability in the data.**

**Because the effect sizes we found in our study were small we did not go on to explore t**he number of subsequent hospitalisations/referrals for depression/cardiovascular disease, or deaths amongst those visiting primary/secondary care for depression/cardiovascular disease in the 1-, 2- and 4-week periods before and after the clock changes.

### ****Text S3: Definitions of outcomes****

**Search and selection of disease and prescription codes**

Medical codes (symptoms/diagnoses) and product codes (prescriptions) were used to identify events in primary care. ICD-10 codes (symptoms/diagnoses) were used to identify events in Hospital Episode Statistics Admitted Patient Care, and A&E codes were used to identify events in Hospital Episode Statistics Accident and Emergency data.

Where available, existing published code lists were used as a starting point for my code lists. Where previous code lists did not exist (e.g. road traffic injuries (ICD-10 & medical codes), sleep (ICD-10 codes)), these were created by searching the ICD-10 dictionary (International Classification of Diseases, 10^th^ revision) or the CPRD GOLD medical dictionary using keywords. All accident and emergency (A&E) code lists were created from scratch using the dataset’s unique coding system. All code lists were checked by a general practitioner experienced in using the codes in clinical practice and refined following their feedback.

For the eight health conditions, codes not relevant to a current diagnosis or symptom (e.g. condition in remission, history of condition) were excluded, as were codes where the timing of the code was not considered to be directly related to the onset of disease (e.g. monitoring/management plans/referrals/advice/counselling). Codes were also excluded if they represented tests (e.g. imaging or sleep/mood rating scales) as they only show a test has been carried out, not that a diagnosis was confirmed.

The HES Accident & Emergency dataset has its own coding system. Road traffic injuries and self-harm were able to be identified on the basis of the ‘aepatgroup’ code which gives the reason for the A&E episode. CVD events were identified on the basis of the ‘aepatgroup’ combined with a 2 digit diagnosis code. It was not possible to specify individual codes for depression, anxiety, eating disorders or sleep disorders. Instead we included the broad category of ‘psychiatric conditions’ in A&E only as a separate condition.

**Anxiety**

Anxiety events in primary and secondary care were defined using ICD-10 and medical codes. ICD-10 codes alone were used to define anxiety. Medical codes alone were used to define anxiety if the code was specific enough. However, where medical codes related to symptoms of anxiety that were not considered adequate to define anxiety, patients were only counted as having anxiety if they were prescribed a drug used to treat anxiety within 90 days either side of the medical code.

ICD-10 codes for the following conditions were included/excluded from our definition:

| **Included** | **Excluded** |
| --- | --- |
| Phobic anxiety disorders  Other anxiety disorders*  Obsessive-compulsive disorder  Reaction to severe stress, and adjustment disorders  Dissociative [conversion] disorders  Other neurotic disorders | Specific (isolated) phobias (e.g. heights)  Somatoform phobias |

*This includes mixed anxiety & depressive disorder which is included in the definitions of both depression and anxiety.

Medical codes for the following conditions (diagnoses & symptoms) were included/excluded from our definition:

| **Included** | **Excluded** |
| --- | --- |
| Generalised anxiety disorder  Panic disorder  Agoraphobia  Social anxiety disorder  Mixed anxiety and depression*  Obsessive compulsive disorders **  Trauma- and stress-related disorders** with anxiety, including PTSD, acute stress disorder, and adjustment disorder with anxiety | Specific phobias (e.g. heights)  Somatic symptoms disorder |

*Mixed anxiety & depressive disorder is included in the definitions of both depression and anxiety.

**Prior to the publication of DSM-5 in 2013, obsessive-compulsive and stress-related disorders were classified as anxiety disorders. In a major change, DSM-5 classified these conditions separately from anxiety disorders. This study uses patient data from 2008-2019. It is not clear how, or whether, the change affected the allocation of Read codes by GPs. The accuracy of the medical codes to identify these conditions may therefore be suboptimal.

Drugs used to treat anxiety:

**Substance name**

Alprazolam

Amitriptyline hydrochloride/ Chlordiazepoxide

Buspirone hydrochloride

Chlordiazepoxide hydrochloride

Diazepam

Duloxetine hydrochloride

Escitalopram oxalate

Lorazepam

Meprobamate

Moclobemide

Oxazepam

Oxprenolol hydrochloride

Paroxetine hydrochloride

Pericyazine

Perphenazine

Pregabalin

Trazodone hydrochloride

Venlafaxine hydrochloride

**Cardiovascular disease (major, acute)**

Major, acute cardiovascular disease events in primary and secondary care were defined with medical codes, ICD-10 and A&E codes.

ICD-10 and medical codes (diagnoses & symptoms) for the following conditions were included/excluded from our definition:

| **Included** | **Excluded** |
| --- | --- |
| Unstable angina/acute coronary syndrome  Myocardial infarction*  Acute ischaemic heart disease  Heart failure  Cardiac arrest  Atrial fibrillation & arrhythmias  Stroke  Transient ischaemic attack |  |

*Codes for events following or subsequent to MI have been included as there is thought to be a high likelihood of miscoding with these MI codes which could lead to MIs being missed.

To be included as an A&E case of major, acute cardiovascular disease events had to have one of the following ‘aepatgroup’ codes (the reason for the A&E visit) and one of the following ‘diag2’ codes (diagnosis codes):

| **aepatgroup** | **diag2** |
| --- | --- |
| Brought in dead  Other than above  Not known | Cardiac conditions  Cerebro-vascular conditions  Other vascular conditions |

**Depression**

Depression events were defined in primary and secondary care using ICD-10 and medical codes. ICD-10 codes alone were used to define depression. Medical codes alone were used to define depression if the code was specific enough. However, where medical codes related to symptoms of depression that were not considered adequate to define depression, patients were only counted as having depression if they were also prescribed a drug used to treat depression within 90 days either side of the medical code.

ICD-10 codes for the following conditions were included/excluded from our definition:

| **Included** | **Excluded** |
| --- | --- |
| Depressive episode  Recurrent depressive disorder  Persistent mood [affective] disorders  Mixed anxiety and depressive disorder*  Depressive conduct disorder |  |

*Mixed anxiety & depressive disorder is included in the definitions of both depression and anxiety.

Medical codes for the following conditions (diagnoses & symptoms) were included/excluded from our definition:

| **Included** | **Excluded** |
| --- | --- |
| Major depressive disorder  Dysthymia  Recurrent depressive disorder  Mixed anxiety and depression*  Disruptive mood dysregulation disorder  Depression in dementia (or other condition)  Trauma- and stress-related disorders with depressed mood (including adjustment disorders with depressed mood**) | Bipolar and related disorders (including bipolar I, II and cyclothymic disorder)  Premenstrual dysphoric disorder  Suicide***  Self-harm***  Maternal depression |

*Mixed anxiety & depressive disorder is included in the definitions of both depression and anxiety.

**Symptoms and treatment of adjustment disorders overlap with both depression and anxiety. To avoid misclassification, adjustment disorders will be included within our definitions of both depression and anxiety. Also see note on stress-related disorders on anxiety table.

***Suicide and self-harm are examined as a separate condition to depression.

Drugs used to treat depression:

**Substance name**

Agomelatine

Amitriptyline Hydrochloride

Amitriptyline Hydrochloride/ Perphenazine

Citalopram hydrobromide

Citalopram hydrochloride

Clomipramine hydrochloride

Dosulepin hydrochloride

Dosulepin Hydrochloride

Duloxetine hydrochloride

Escitalopram oxalate

Fluoxetine hydrochloride

Fluvoxamine maleate

Imipramine hydrochloride

Isocarboxazid

Lofepramine hydrochloride

Mianserin hydrochloride

Mirtazapine

Moclobemide

Nortriptyline hydrochloride

Nortriptyline Hydrochloride

Paroxetine hydrochloride

Phenelzine sulfate

Reboxetine mesilate

Sertraline

Sertraline hydrochloride

Tranylcypromine sulfate

Trazodone Hydrochloride

Trimipramine maleate

Venlafaxine hydrochloride

Venlafaxine Hydrochloride

Vortioxetine hydrobromide

**Eating disorders**

Eating disorder events in primary and secondary care were defined with ICD-10 and medical codes. Because we are looking at the effect of the clock changes (via sleep) on eating disorders we excluded organic conditions which are caused by an underlying medical condition rather than having a psychiatric cause.

ICD-10 codes for the following conditions were included/excluded from our definition:

| **Included** | **Excluded** |
| --- | --- |
| Anorexia nervosa (including atypical)  Bulimia nervosa (including atypical)  Overeating/vomiting associated with psychological disturbances  Unspecified/other eating disorder |  |

Medical codes for the following conditions were included/excluded from our definition:

| **Included (diagnoses & symptoms)** | **Excluded** |
| --- | --- |
| Anorexia nervosa (including atypical)  Bulimia nervosa  Compulsive eating disorder  Non-organic* eating disorders (specified or unspecified)  Hyperorexia nervosa  Overeating associated with psychological disturbances/  psychogenic overeating  Other eating disorders (including unspecified)  Nocturnal sleep-related eating disorder | Organic eating disorders (without an overt psychiatric cause) e.g. hyperalimentation, pica, starvation, anorexia (without nervosa suffix), excessive eating/polyphagia without psychological disturbance. |

*Nonorganic eating disorders are those where emotional/psychiatric causes are believed to be the main contributory factor rather than organic eating disorders which are caused by an underlying physical medical condition.

**Psychiatric conditions**

It was not possible to specify individual codes for anxiety, depression, eating disorders or sleep disorders in the A&E data. Instead we examined the broader condition of ‘psychiatric conditions’. To be included as a psychiatric condition in A&E, events had to have one of the following ‘aepatgroup’ codes (the reason for the A&E visit) and the following ‘diag2’ code (diagnosis code):

| **aepatgroup** | **diag2** |
| --- | --- |
| Brought in dead  Other than above  Not known | Psychiatric conditions |

**Road traffic injuries**

Road traffic injury events in primary and secondary care were defined with medical codes, ICD-10 and A&E codes.

ICD-10 codes for the following conditions were included/excluded from our definition:

| **Included** | **Excluded** |
| --- | --- |
| Land traffic accident involving a motor vehicle  Unspecified / other specified land transport accident  Unspecified vehicle land transport accident  Crashing of motor vehicle, undetermined intent | Land transport accidents specifically reported as ‘non-traffic’.  Accidents specifically reporting not involving motor vehicles (e.g. pedestrian hit by pedal bike/train)  Accidents to persons engaged in the maintenance or repair of transport equipment or vehicle (not in motion) unless injured by another vehicle in motion  Accidents involving vehicles, but unrelated to the hazards associated with the means of transportation, e.g., injuries received in a fight on board ship; transport vehicle involved in a cataclysm; finger crushed when shutting car door  Intentional self-harm* |

*Suicide and self-harm are examined as a separate condition to road traffic injuries.

Medical codes for the following conditions were included/excluded from our definition:

| **Included** | **Excluded** |
| --- | --- |
| Traffic accidents involving motor vehicles (including MVTA*)  Crashing of motor vehicle undetermined intent  Road vehicle accidents not otherwise specified  Other road vehicle accidents  RTA – road traffic & other transport accidents  Accidents involving vehicles not elsewhere classifiable/not otherwise specified  Vehicle accident not elsewhere classifiable/not otherwise specified | Transport accidents that specifically report being ‘non-traffic’ (including (MVNTA**)  Accidents that specifically report not involving motor vehicles (e.g. pedestrian hit by non-motor vehicle or pedal bike)  Intentional self-harm/ suicide/jumping in front of vehicle/exhaust gas poisoning*** |

*MVTA = motor vehicle traffic accident

**MVNTA = motor vehicle non-traffic accident

***Suicide and self-harm are examined as a separate condition to road traffic injuries.

To be included as a road traffic injury in A&E, events had to have the following ‘aepatgroup’ code (the reason for the A&E visit):

| **aepatgroup** |
| --- |
| Road traffic accident |

**Self-harm**

Intentional self-harm events in primary and secondary care were defined using ICD-10, medical codes and A&E codes.

ICD-10 codes for the following conditions were included/excluded from our definition:

| **Included** | **Excluded** |
| --- | --- |
| Intentional self-harm  Sequelae of intentional self-harm | Events of undetermined intent  Sequelae of events undetermined intent |

Medical codes for the following conditions (diagnoses & symptoms) were included/excluded from our definition:

| **Included** | **Excluded** |
| --- | --- |
| Suicide/attempted suicide  Self-harm (intentional, self-inflected injury, poisoning, overdose through any means) | Events of undetermined intent  Accidental injury/self-harm  Events undetermined whether self-inflicted |

To be included as self-harm A&E, events had to have the following ‘aepatgroup’ code (the reason for the A&E visit):

| **aepatgroup** |
| --- |
| Deliberate self-harm |

**Sleep disorders**

Sleep disorder events in primary and secondary care were defined using ICD-10 and medical codes. ICD-10 codes alone were used to define sleep disorders. Sleep disorders are difficult to define which means that most Read codes are not specific and relate to symptoms rather than diagnoses. Patients were therefore only counted as having a sleep disorder if they had a Read code for a sleep disorder diagnosis or symptom and were also prescribed a drug used to treat sleep disorders within 90 days either side of the medical code.

ICD-10 codes for the following conditions were included/excluded from our definition:

| **Included** | **Excluded** |
| --- | --- |
| Disorders of initiating and maintaining sleep [insomnias]  Disorders of excessive somnolence [hypersomnias]  Disorders of the sleep-wake schedule  Other/unspecified sleep disorders/disorders  Nonorganic* insomnia/hypersomnia/disorders of sleep-wake schedule  Sleep walking/ sleep terrors/nightmares  Other/unspecified nonorganic sleep disorders | Sleep apnoea  Narcolepsy  Cataplexy |

*Nonorganic sleep disorders are those where emotional causes are believed to be the main contributory factor rather than an underlying physical medical condition.

Medical codes for the following conditions (diagnoses & symptoms) were included/excluded from our definition:

| **Included** | **Excluded** |
| --- | --- |
| Insomnia  Hypersomnia  Circadian rhythm sleep-wake disorders  Paraomnias  Sleep quality/insomnia indexes | Narcolepsy  Breathing-related disorders (including sleep apnoea)  Cataplexy |

Drug substances used to treat sleep disorders:

| **Included** | **Excluded*** |
| --- | --- |
| Temazepam | Promethazine hydrochloride |
| Diazepam | Promethazine hydrochloride/pholcodine |
| Zopiclone | Pethidine hydrochloride/promethazine hydrochloride |
| Clomethiazole | Paracetamol/promethazine hydrochloride/ dextromethorphan hydrobromide |
| Zolpidem tartrate | Dextromethorphan hydrobromide/promethazine hydrochloride/ paracetamol |
| Clomethiazole edisilate |  |
| Flunitrazepam |  |
| Flurazepam hydrochloride |  |
| Lormetazepam |  |
| Oxazepam |  |
| Loprazolam mesilate |  |
| Melatonin |  |
| Nitrazepam |  |

*We excluded antihistamines, cough syrups and cold & flu relief products that are unlikely to be prescribed for a sleep disorder.

### ****Text S4: Creation of code lists from scratch****

**ICD10 code lists**

There were no existing ICD10 code lists for road traffic injuries, eating disorders or sleep disorders so these were created by manually searching the ICD-10 hierarchy (https://icd.who.int/browse10/2016/en).

**Medical code lists**

There were no existing medical code lists for road traffic injuries. Instead this code list was created by searching descriptions in the CPRD gold medical dictionary (June 2022 build) for the following terms: "*motor*" "*traffic*" "*mvta*" "*vehicle*" "*road*" "*collision*" "*rta*" "*rtc*. Codes not related to road traffic injuries were removed.

### ****Text S5: CODE-EHR Checklist****

CODE-EHR framework: Best practice checklist to report on the use of structured electronic healthcare records in clinical research

Date of completion: 11/Apr/25	Study name: The Effects of Daylight Saving Time Clock Changes on Health in England: A Population Based Retrospective Cohort Study Using Linked Electronic Health Records

| **Item** | **Objective** | **Framework standards** | **Minimum information to provide** | **Lead Author acknowledgement** |
| --- | --- | --- | --- | --- |
| 1. Dataset construction and linkage | To provide an understanding of how the structured healthcare data were identified and used. | Minimum: Flow diagram of datasets used in the study, and description of the processes and directionality of any linkage performed, published within the research report or supplementary documents.  Preferred: Provided within a pre-published protocol or open-access document. | (a) State the source of any datasets used.  (b) Comment on how the observed and any missing data were identified and addressed, and the proportion observed for each variable. (c) Provide data on completeness of follow-up. (d) For linked datasets, specify how linkage was performed and the quality of linkage methods. | Select one option:  Minimum standard not met ☐  Minimum standard met ☐  Preferred standard met ☒ |
| 2. Data fit for purpose | To ensure transparency with the approach taken, with respect to coding of the structured healthcare data. | Minimum: Clear unambiguous statements on the process of coding in the methods section of the research report.  Preferred: Provided within a pre-published protocol or open-access document. | (a) Confirm origin, clinical processes, and the purpose of data. (b) Specify coding systems, clinical terminologies or classification used and their versions, and any manipulation of the coded data.  (c) Provide detail on quality assessment for data capture. (d) Outline potential sources of bias. | Select one option:  Minimum standard not met ☐  Minimum standard met ☐  Preferred standard met ☒ |
| 3. Disease and outcome definitions | To fully detail how conditions AND outcome events were defined, allowing other researchers to identify errors and repeat the process in other datasets. | Minimum: State what codes were used to define diseases, treatments, conditions and outcomes *prior to statistical analysis*, including those relating to patient identification, therapy, procedures, comorbidities, and components of any composite endpoints.  Preferred: Provided within a pre-published protocol or open-access document *prior to statistical analysis.* | (a) Detailed lists of codes used for each aspect of the study.  (b) Date of publication and access details for the coding manual (please add to box below). (c) Provide definitions, implementation logic and validation of any phenotyping algorithms used. (d) Specify any processes used to validate the coding scheme or reference to prior work. | Select one option:  Minimum standard not met ☐  Minimum standard met ☐  Preferred standard met ☒ |
| 4. Analysis | To fully detail how outcome events were analysed and allow independent assessment of the authenticity of study findings. | Minimum: Describe the process used to analyse study outcomes, including statistical methods and use of any machine learning or algorithmic approaches.  Preferred: Provide a statistical analysis plan as a supplementary file, locked *prior to* *analyses commencing.* | (a) Provide details on all statistical methods used. (b) Provide links to any machine code or algorithms used in the analysis, preferably as open-source. (c) Specify the processes of testing assumptions, assessing model fit and any internal validation.  (d) Specify how generalisability of results was assessed, the replication of findings in other datasets, or any external validation. | Select one option:  Minimum standard not met ☐  Minimum standard met ☐  Preferred standard met ☒ |
| 5. Ethics and governance | To provide patients, who may or may not have given consent, and regulatory authorities the ability to interrogate the security and provenance of the data. | Minimum: Clear unambiguous statements on how the principles of Good Clinical Practice and Data Protection will be/were met, provided in the methods section of the research report.  Preferred: Provided within a pre-published protocol or open-access document with evidence of patient and public engagement. | (a) State how informed consent was acquired, or governance if no patient consent. (b) Specify how data privacy was protected in the collection and storage of data.  (c) Detail what steps were taken for patient and public involvement in the research study. (d) Provide information on where anonymised source data or code can be obtained for verification and further research. | Select one option:  Minimum standard not met ☐  Minimum standard met ☒  Preferred standard met ☐ |
| 6. Coding manual | DOI of publication or website address: Click or tap here to enter text.  Date published: Click to enter a date | | | |
| 7. Comments | Click or tap here to enter text. | | | |
| **8. Summary declaration** | One or more minimum standards not met ☐ OR **All minimum standards met** ☒  Number of preferred standards met: 4 / 5 | | | |

### ****Text S6: STROBE-RECORD Statement****

**The RECORD statement – checklist of items, extended from the STROBE statement, that should be reported in observational studies using routinely collected health data.**

|  | **Item No.** | **STROBE items** | **Location in manuscript where items are reported** | **RECORD items** | **Location in manuscript where items are reported** |
| --- | --- | --- | --- | --- | --- |
| **Title and abstract** | | |  |  |  |
|  | 1 | (a) Indicate the study’s design with a commonly used term in the title or the abstract (b) Provide in the abstract an informative and balanced summary of what was done and what was found |  | RECORD 1.1: The type of data used should be specified in the title or abstract. When possible, the name of the databases used should be included.    RECORD 1.2: If applicable, the geographic region and timeframe within which the study took place should be reported in the title or abstract.    RECORD 1.3: If linkage between databases was conducted for the study, this should be clearly stated in the title or abstract. | Subtitle Title & Abstract  Title &  Abstract  Abstract |
| **Introduction** | | |  |  |  |
| Background rationale | 2 | Explain the scientific  background and rationale for the investigation being reported | Introduction |  |  |
| Objectives | 3 | State specific objectives, including any prespecified hypotheses | Introduction |  |  |
| **Methods** | | |  |  |  |
| Study Design | 4 | Present key elements of study design early in the paper | Abstract  Method: Data resource and study population |  |  |
| Setting | 5 | Describe the setting, locations, and relevant dates, including periods of recruitment, exposure, follow-up, and data collection | Abstract.  Method: Data resource, study population, exposures, outcomes, covariates. |  |  |

| Participants | 6 | (a) *Cohort study* - Give the eligibility criteria, and the sources and methods of selection of participants. Describe methods of follow-up   *Case-control study* - Give the eligibility criteria, and the sources and methods of case ascertainment and control selection. Give the rationale for the choice of cases and controls *Cross-sectional study* - Give the eligibility criteria, and the sources and methods of selection of participants     (b) *Cohort study* - For matched studies, give matching criteria and number of exposed and unexposed   *Case-control study* - For matched studies, give matching criteria and the number of controls per case |  | RECORD 6.1: The methods of study population selection (such as codes or algorithms used to identify subjects) should be listed in detail. If this is not possible, an explanation should be provided.    RECORD 6.2: Any validation studies of the codes or algorithms used to select the population should be referenced. If validation was conducted for this study and not published elsewhere, detailed methods and results should be provided.    RECORD 6.3: If the study involved linkage of databases, consider use of a flow diagram or other graphical display to demonstrate the data linkage process, including the number of individuals with linked data at each stage. | Method, paragraph 3 & Figure 1. Supplementary text S3 & S4. Code lists on  Github repository.  N/A  Method, Study population |
| --- | --- | --- | --- | --- | --- |
| Variables | 7 | Clearly define all outcomes, exposures, predictors, potential confounders, and effect modifiers. Give diagnostic criteria, if applicable. |  | RECORD 7.1: A complete list of codes and algorithms used to classify exposures, outcomes, confounders, and effect modifiers should be provided. If these cannot be reported, an explanation should be provided. | Supplementary text S4 & Github repository. |
| Data sources/ measurement | 8 | For each variable of interest, give sources of data and details of methods of assessment (measurement).  Describe comparability of assessment methods if there is  more than one group | Method, supplementary tables S1-3, supplementary text S3 & code lists on GitHub. |  | . |

| Bias | 9 | Describe any efforts to address potential sources of bias | Sensitivity analyses to explore the effects of individuals having multiple events over an 8-week clock change period, described in Statistical analysis of Method section. |  |  |
| --- | --- | --- | --- | --- | --- |
| Study size | 10 | Explain how the study size was arrived at | Method: Study Population section |  |  |
| Quantitative variables | 11 | Explain how quantitative variables were handled in the analyses. If applicable, describe which groupings were chosen,  and why | Method: outcomes, covariates, statistical analysis & supplementary text S2. |  |  |
| Statistical methods | 12 | (a) Describe all statistical methods, including those used to control for confounding (b) Describe any methods used to examine subgroups and interactions   (c) Explain how missing data were addressed (d) *Cohort study* - If applicable, explain how loss to follow-up was addressed   *Case-control study* - If applicable, explain how matching of cases and controls was addressed  *Cross-sectional study* - If applicable, describe analytical methods taking account of sampling strategy   (e) Describe any sensitivity analyses | Method, Statistical Analysis section |  |  |
| Data access and cleaning methods |  | .. |  | RECORD 12.1: Authors should describe the extent to which the investigators had access to the database population used to create the study population. | Method, study population section. |

|  |  |  |  | RECORD 12.2: Authors should provide information on the data cleaning methods used in the study. | Method - Study population, outcomes & covariates sections. Supplementary table S2. |
| --- | --- | --- | --- | --- | --- |
| Linkage |  | .. |  | RECORD 12.3: State whether the study included person-level,  institutional-level, or other data linkage across two or more databases. The methods of linkage and methods of linkage quality evaluation should be provided. | Method: Study population section. |
| **Results** | | | | | |
| Participants | 13 | (a) Report the numbers of individuals at each stage of the study (*e.g.*, numbers potentially eligible, examined for eligibility, confirmed eligible, included in the study, completing follow-up, and analysed) (b) Give reasons for nonparticipation at each stage. (c) Consider use of a flow diagram |  | RECORD 13.1: Describe in detail the selection of the persons included in the study (*i.e.,* study population selection) including filtering based on data quality, data availability and linkage. The selection of included persons can be described in the text and/or by means of the study flow diagram. | Method, Study population & Figure 1. |
| Descriptive data | 14 | (a) Give characteristics of study participants (*e.g.*, demographic, clinical, social) and information on exposures and potential   confounders   (b) Indicate the number of participants with missing data for each variable of interest (c) *Cohort study* - summarise follow-up time (*e.g.*, average and total amount) | Table 1 |  |  |
| Outcome data | 15 | *Cohort study* - Report numbers of outcome events or summary measures over time  *Case-control study* - Report numbers in each exposure  category, or summary measures of exposure  *Cross-sectional study* - Report numbers of outcome events or summary measures | Figure 1. |  |  |
| Main results | 16 | (a) Give unadjusted estimates and, if applicable, confounderadjusted estimates and their precision (e.g., 95% confidence interval). Make clear which confounders were adjusted for and why they were included (b) Report category boundaries when continuous variables were categorized  (c) If relevant, consider translating estimates of relative risk into absolute risk for a meaningful time period | Results, table 2 & supplementary tables S4-S7. |  |  |
| Other analyses | 17 | Report other analyses done—  e.g., analyses of subgroups and interactions, and sensitivity analyses | Results: Events in the first week after the clock changes section & negative exposure section. Supplementary tables S4-S8. |  |  |
| **Discussion** | | | | | |
| Key results | 18 | Summarise key results with reference to study objectives | Discussion, paragraph 1. |  |  |
| Limitations | 19 | Discuss limitations of the study, taking into account sources of potential bias or imprecision. Discuss both direction and magnitude of any potential bias |  | RECORD 19.1: Discuss the  implications of using data that were not created or collected to answer the specific research question(s). Include discussion of misclassification bias, unmeasured confounding, missing data, and changing eligibility over time, as they pertain to the study being reported. | Discussion (paragraphs 7-8) |
| Interpretation | 20 | Give a cautious overall interpretation of results considering objectives,  limitations, multiplicity of analyses, results from similar studies, and other relevant evidence | Discussion, paragraphs 10-13 |  |  |
| Generalisability | 21 | Discuss the generalisability (external validity) of the study results | N/A |  |  |
| **Other Information** | | | | | |
| Funding | 22 | Give the source of funding and the role of the funders for the present study and, if applicable, for the original study on which the present article is based | Funding statement |  |  |
| Accessibility of protocol, raw data, and programming code |  | .. |  | RECORD 22.1: Authors should provide information on how to access any supplemental information such as the study protocol, raw data, or programming code. | Supplementary text (including protocol), tables & figures provided & referenced in the paper. Data sharing statement provided including link to GitHub repository with full code. |

*Reference: Benchimol EI, Smeeth L, Guttmann A, Harron K, Moher D, Petersen I, Sørensen HT, von Elm E, Langan SM, the RECORD Working Committee. The REporting of studies Conducted using Observational Routinely-collected health Data (RECORD) Statement. *PLoS Medicine* 2015; in press.

*Checklist is protected under Creative Commons Attribution ([CC BY)](http://creativecommons.org/licenses/by/4.0/) license.

### Table S1: List of dates of DST clock changes 2008-2019

| **Year** | **Date of spring clock change** | **Date of autumn clock change** |
| --- | --- | --- |
| 2008 | Sunday 30^th^ March | Sunday 26^th^ October |
| 2009 | Sunday 29^th^ March | Sunday 25^th^ October |
| 2010 | Sunday 28^th^ March | Sunday 31^st^ October |
| 2011 | Sunday 27^th^ March | Sunday 30^th^ October |
| 2012 | Sunday 25^th^ March | Sunday 28^th^ October |
| 2013 | Sunday 31^st^ March | Sunday 27^th^ October |
| 2014 | Sunday 30^th^ March | Sunday 26^th^ October |
| 2015 | Sunday 29^th^ March | Sunday 25^th^ October |
| 2016 | Sunday 27^th^ March | Sunday 30^th^ October |
| 2017 | Sunday 26^th^ March | Sunday 29^th^ October |
| 2018 | Sunday 25^th^ March | Sunday 28^th^ October |
| 2019 | Sunday 31^st^ March | Sunday 27^th^ October |

### ****Table S2: Derivation of covariates****

| **Covariate** | **Categories** | **Derivation** |
| --- | --- | --- |
| Age | Anxiety: 10-50, >50  CVD: 40-75, >75  Depression: 10-45, >45  Eating disorders: 10-25, >25  Psychiatric conditions in A&E: 10-35, >35  Road traffic injuries: 0-30, >30  Self-harm: 10-30, >30  Sleep: 10-55, >55 | Recorded in primary care record.  To maximise statistical power we created binary variables for age at the time of the clock change, with roughly equally-sized groups based on the median. The age categories were therefore different for each health condition.  CPRD does not collect exact date of birth due to the date needing to be anonymised. All patients without a day of birth had this inputted as 1. All participants without a month of birth (those aged 16 & over) had month inputted as July. |
| Sex | Male, Female, missing | Recorded in primary care record. ‘Data not entered’, ‘indeterminate’ or ‘unknown’ were categorised as missing. However, the missing category was not included in stratified results tables due to sample sizes being too small. |
| Marital status | In relationship, not in relationship, missing | Recorded in primary care record.  ‘Married’, ‘engaged’, ‘co-habiting’, ‘re-married’, ‘stable relationship’, ‘civil partnership’ recategorised as ‘in relationship’.  ‘Single’, ‘widowed’, ‘divorced’, ‘separated’ categorised as ‘not in relationship’.  ‘Data not entered’, ‘unknown’ categorised as missing. |
| Incident vs prevalent case | Yes, no | First ever event for this condition recorded in either primary or secondary care vs previous event for this condition recorded in either primary or secondary care. |
| Deprivation | Most deprived, rest, missing | Patient-level Index of Multiple Deprivation for England (2019) based on patient postcode. Provided by CPRD through third-party linkage to Office for National Statistics data.  20 categories recategorised to create a binary variable with the most deprived 20% included in the ‘most deprived’ category.  The missing category was not included in stratified results tables due to sample sizes being too small. |
| BMI | Overweight/obese, normal/underweight, missing | Recorded in primary care record. Defined as the most recent record prior to the clock change.  Extreme values (BMI <12 or >50 kg/m^2^) were excluded. Remaining values recategorised: Overweight/obese: ≥ 25 kg/m^2^, normal/underweight <25 kg/m^2^.  <https://www.nhs.uk/conditions/obesity/> |
| Alcohol status | Current drinker, non/ex-drinker, missing | Recorded in primary care record. Defined as the most recent record prior to the clock change.  ‘Non-drinker’ and ‘ex-drinker’ combined to create ‘non/ex-smoker’ category. |
| Smoking status | Current smoker, non/ex-smoker, missing | Recorded in primary care record. Defined as the most recent record prior to the clock change.  ‘Non-smoker’ and ‘ex-smoker’ combined to create ‘non/ex-smoker’ category. |
| Systolic blood pressure | High, normal, missing | Recorded in primary care record. Defined as the most recent record prior to the clock change.  Raw values recategorised to be ‘High’: ≥140 mmHg or ‘normal’ <140 mmHg.  <https://www.ncbi.nlm.nih.gov/pmc/articles/PMC381142/> |
| Diastolic blood pressure | High, normal, missing | Recorded in primary care record. Defined as the most recent record prior to the clock change.  Raw values recategorised to be ‘High’: ≥90 mmHg or ‘normal’ <90 mmHg.  <https://www.ncbi.nlm.nih.gov/pmc/articles/PMC381142/> |
| Cardiovascular event subgroup | Atrial fibrillation/arrhythmia, myocardial infarction, stroke/transient ischemic attack, other | Based on Medical code or ICD10 code (primary care and HES APC data only) |
| Easter weekend | Yes, no | Binary variable based on whether the event date fell on one of the 5 days of the Easter weekend (including Maundy Thursday). |
| Region | 9 numeric categories indicating where in England the GP practice is, based on its Strategic Health Authority | Provided by CPRD. Categories: North East, North West, Yorkshire & The Humber, East Midlands, West Midlands, East of England, London, South East and South West |

### Table S3: List of dates of 5-day Easter weekends 2008-2019

| **Year** | **Easter Sunday** | **Easter weekend start/end** |
| --- | --- | --- |
| 2008 | 23^rd^ March | 20^th^-24^th^ March |
| 2009 | 12^th^ April | 9^th^-13^th^ April |
| 2010 | 4^th^ April | 1^st^-5^th^ April |
| 2011 | 24^th^ April | 21^st^-25^th^ April |
| 2012 | 8^th^ April | 5^th^-9^th^ April |
| 2013 | 31^st^ March | 28^th^ March-1^st^ April |
| 2014 | 20^th^ April | 17^th^-21^st^ April |
| 2015 | 5^th^ April | 2^nd^-6^th^ April |
| 2016 | 27^th^ March | 24^th^-28^th^ April |
| 2017 | 16^th^ April | 13^th^-17^th^ April |
| 2018 | 1^st^ April | 29^th^ March-2^nd^ April |
| 2019 | 21^st^ April | 18^th^-22^nd^ April |

### Table S4: Adjusted* rate ratios of the number of events in the 1 week (including by day and Monday-Friday), 2 week and 4 week periods after the Spring and Autumn clock changes (England, 2008-2019) versus control periods**

| **Anxiety** | | | | | | |
| --- | --- | --- | --- | --- | --- | --- |
|  | **Spring** | | | **Autumn** | | |
|  | **Events** | **IRR (95% CI)** | **p value** | **Events** | **IRR (95% CI)** | **p value** |
| Sunday | 3,869 | 0.94 (0.802, 1.092) | 0.3997 | 4,468 | 0.98 (0.888, 1.072) | 0.6114 |
| Monday | 18,008 | 1.00 (0.941, 1.065) | 0.9780 | 20,600 | 0.94 (0.904, 0.986) | 0.0095 |
| Tuesday | 18,324 | 1.00 (0.959, 1.034) | 0.8269 | 18,956 | 0.96 (0.912, 1.018) | 0.1832 |
| Wednesday | 17,062 | 1.00 (0.954, 1.044) | 0.9303 | 18,229 | 0.96 (0.931, 0.999) | 0.0460 |
| Thursday | 17,256 | 0.99 (0.933, 1.053) | 0.7820 | 18,408 | 0.97 (0.942, 0.997) | 0.0326 |
| Friday | 15,517 | 1.00 (0.950, 1.062) | 0.8722 | 17,909 | 0.95 (0.915, 0.983) | 0.0037 |
| Saturday | 4,892 | 0.97 (0.898, 1.038) | 0.3395 | 5,434 | 1.06 (0.949, 1.189) | 0.2917 |
| Mon-Fri | 86,167 | 1.00 (0.970, 1.041) | 0.7904 | 94,102 | 0.96 (0.942, 0.974) | 7.133x10^-07^ |
| 1 week | 94,928 | 0.99 (0.958, 1.029) | 0.6954 | 104,004 | 0.97 (0.947, 0.984) | 0.0002 |
| 2 weeks | 94,928 | 0.98 (0.960, 0.995) | 0.0111 | 104,004 | 1.01 (0.996, 1.019) | 0.2073 |
| 4 weeks | 94,928 | 1.00 (0.982, 1.013) | 0.7492 | 104,004 | 1.01 (0.997, 1.027) | 0.1135 |
| **Acute cardiovascular disease** | | | | | | |
|  | **Spring** | **Autumn** |  |  |  |  |
|  | **Events** | **IRR (95% CI)** | **P value** | **Events** | **IRR (95% CI)** | **P value** |
| Sunday | 27,671 | 1.00 (0.962, 1.034) | 0.8902 | 27,275 | 0.97 (0.905, 1.032) | 0.3130 |
| Monday | 48,591 | 1.02 (0.978, 1.055) | 0.4312 | 50,109 | 0.95 (0.916, 0.983) | 0.0039 |
| Tuesday | 49,904 | 1.03 (0.987, 1.071) | 0.1774 | 49,640 | 1.00 (0.963, 1.036) | 0.9494 |
| Wednesday | 49,020 | 1.04 (1.003, 1.077) | 0.0324 | 48,232 | 0.97 (0.940, 1.003) | 0.0770 |
| Thursday | 49.024 | 1.00 (0.953, 1.053) | 0.9381 | 48,454 | 0.95 (0.914, 0.990) | 0.0132 |
| Friday | 45,443 | 1.02 (0.979, 1.061) | 0.3594 | 48,007 | 0.97 (0.936, 1.001) | 0.0592 |
| Saturday | 29,150 | 1.01 (0.979, 1.044) | 0.5044 | 29,792 | 1.05 (1.004, 1.107) | 0.0328 |
| Mon-Fri | 241,982 | 1.02 (1.008, 1.038) | 0.0026 | 244,442 | 0.97 (0.946, 0.990) | 0.0046 |
| 1 week | 298,803 | 1.02 (1.005, 1.030) | 0.0058 | 301,509 | 0.98 (0.958, 0.999) | 0.0423 |
| 2 weeks | 298,803 | 1.00 (0.991, 1.015) | 0.6558 | 301,509 | 1.00 (0.981, 1.015) | 0.8351 |
| 4 weeks | 298,803 | 0.99 (0.983, 1.001) | 0.0747 | 301,509 | 1.01 (1.000, 1.0170 | 0.0505 |
| **Depression** | | | | | | |
|  | **Spring** | | | **Autumn** | |  |
|  | **Events** | **IRR (95% CI)** | **p value** | **Events** | **IRR (95% CI)** | **p value** |
| Sunday | 7,118 | 0.92 (0.867, 0.984) | 0.0143 | 7,807 | 0.95 (0.898, 1.006) | 0.0780 |
| Monday | 50,837 | 0.99 (0.956, 1.016) | 0.3344 | 55,831 | 0.93 (0.905, 0.963) | 1.593x10^-5^ |
| Tuesday | 51,149 | 0.98 (0.957, 0.998) | 0.0326 | 51,149 | 0.96 (0.927, 1.001) | 0.0561 |
| Wednesday | 48,860 | 1.00 (0.958, 1.037) | 0.8621 | 48,615 | 0.97 (0.938, 0.995) | 0.0203 |
| Thursday | 48,395 | 1.01 (0.965, 1.048) | 0.8018 | 47,986 | 0.96 (0.926, 0.985) | 0.0039 |
| Friday | 42,959 | 1.01 (0.976, 1.044) | 0.5791 | 47,688 | 0.96 (0.916, 1.007) | 0.0927 |
| Saturday | 9,113 | 1.03 (0.980, 1.089) | 0.2234 | 9,989 | 0.99 (0.920, 1.067) | 0.7990 |
| Mon-Fri | 242,200 | 1.00 (0.983, 1.020) | 0.8759 | 251,269 | 0.96 (0.941, 0.970) | 6.080x10^-09^ |
| 1 week | 258,431 | 1.00 (0.978, 1.012) | 0.5765 | 269,065 | 0.96 (0.946, 0.970) | 9.080x10^-11^ |
| 2 weeks | 258,431 | 0.98 (0.971, 0.994) | 0.0033 | 269,065 | 0.98 (0.966, 0.991) | 0.0010 |
| 4 weeks | 258,431 | 0.99 (0.979, 1.007) | 0.3131 | 269,065 | 1.00 (0.983, 1.009) | 0.5217 |
| **Eating disorders** | | | | | | |
|  | **Spring** | | | **Autumn** | | |
|  | **Events** | **IRR (95% CI)** | **p value** | **Events** | **IRR (95% CI)** | **p value** |
| Sunday | 134 | 0.87 (0.392, 1.913) | 0.7221 | 153 | 0.82 (0.532,1.262) | 0.3656 |
| Monday | 545 | 0.99 (0.726, 1.340) | 0.9284 | 531 | 0.76 (0.566, 1.032) | 0.0790 |
| Tuesday | 486 | 0.99 (0.724, 1.348) | 0.9395 | 463 | 0.64 (0.496, 0.833) | 0.0008 |
| Wednesday | 460 | 1.09 (0.895, 1.319) | 0.4009 | 459 | 1.01 (0.722, 1.422) | 0.9380 |
| Thursday | 525 | 1.15 (0.873, 1.510) | 0.3230 | 488 | 1.49 (1.128, 1.960) | 0.0048 |
| Friday | 413 | 0.76 (0.516, 1.123) | 0.1695 | 477 | 1.10 (0.912, 1.331) | 0.3145 |
| Saturday | 172 | 0.92 (0.558, 1.501) | 0.7254 | 183 | 0.94 (0.522, 1.706) | 0.8478 |
| Mon-Fri | 2,429 | 1.01 (0.933, 1.094) | 0.8023 | 2,418 | 0.99 (0.846, 1.161) | 0.9069 |
| 1 week | 2,735 | 1.00 (0.906, 1.104) | 0.9984 | 2,754 | 0.97 (0.878, 1.082) | 0.6293 |
| 2 weeks | 2,735 | 0.97 (0.912, 1.033) | 0.3465 | 2,754 | 0.99 (0.933, 1.045) | 0.6601 |
| 4 weeks | 2,735 | 1.04 (0.939, 1.147) | 0.4672 | 2,754 | 0.98 (0.937, 1.032) | 0.4983 |
| **Psychiatric conditions (in A&E only)** | | | | | | |
|  | **Spring** | | | **Autumn** | | |
|  | **Events** | **IRR (95% CI)** | **p value** | **Events** | **IRR (95% CI)** | **p value** |
| Sunday | 2695 | 0.92 (0.803, 1.060) | 0.2546 | 2,977 | 0.90 (0.843, 0.962) | 0.0018 |
| Monday | 2893 | 1.11 (0.968, 1.269) | 0.1352 | 3,203 | 0.90 (0.793, 1.012) | 0.0776 |
| Tuesday | 2814 | 0.97 (0.881, 1.078) | 0.6151 | 3,106 | 0.93 (0.837, 1.026) | 0.1450 |
| Wednesday | 2842 | 0.90 (0.750, 1.077) | 0.2469 | 2,967 | 0.98 (0.878, 1.083) | 0.6379 |
| Thursday | 2725 | 0.97 (0.892, 1.054 | 0.4718 | 3,017 | 0.95 (0.884, 1.019) | 0.1477 |
| Friday | 2695 | 1.00 (0.846, 1.181 | 0.9947 | 2,981 | 0.98 (0.862, 1.111) | 0.7373 |
| Saturday | 2604 | 1.05 (0.930, 1.193) | 0.4154 | 2,822 | 0.97 (0.880, 1.075) | 0.5819 |
| Mon-Fri | 13,969 | 0.99 (0.942, 1.035) | 0.6011 | 15,274 | 0.94 (0.891, 1.002) | 0.0579 |
| 1 week | 19,268 | 0.99 (0.932, 1.044) | 0.6289 | 21,073 | 0.94 (0.902, 0.984) | 0.0078 |
| 2 weeks | 19,268 | 0.98 (0.944, 1.016) | 0.2745 | 21,073 | 1.00 (0.977, 1.029) | 0.8411 |
| 4 weeks | 19,268 | 1.00 (0.973, 1.034) | 0.8257 | 21,073 | 1.02 (0.976, 1.063) | 0.4049 |
| **Road traffic injuries** | | | | | | |
|  | **Spring** | |  | **Autumn** |  |  |
|  | **Events** | **IRR (95% CI)** | **p value** | **Events** | **IRR (95% CI)** | **p value** |
| Sunday | 3838 | 1.02 (0.904, 1.147) | 0.7624 | 4,210 | 0.90 (0.791, 1.022) | 0.1036 |
| Monday | 7261 | 0.88 (0.789, 0.992) | 0.0355 | 8,065 | 0.95 (0.839, 1.076) | 0.4208 |
| Tuesday | 6854 | 0.98 (0.899, 1.076) | 0.7226 | 7,250 | 0.87 (0.760, 0.993) | 0.0390 |
| Wednesday | 6589 | 0.97 (0.886, 1.067 | 0.5532 | 7,118 | 0.99 (0.901, 1.093) | 0.8724 |
| Thursday | 6474 | 1.07 (0.966, 1.180) | 0.1979 | 7,107 | 0.98 (0.945, 1.013 | 0.2244 |
| Friday | 6408 | 1.03 (0.924, 1.145) | 0.6054 | 7,406 | 1.09 (0.985, 1.198 | 0.0989 |
| Saturday | 4271 | 0.99 (0.927, 1.054) | 0.7172 | 4,708 | 1.04 (0.921, 1.166) | 0.5528 |
| Mon-Fri | 33,586 | 0.99 (0.940, 1.043) | 0.7069 | 36,946 | 0.97 (0.942, 1.007) | 0.1239 |
| 1 week | 41,695 | 0.99 (0.951, 1.035) | 0.7140 | 45,864 | 0.97 (0.943, 1.003) | 0.0797 |
| 2 weeks | 41,695 | 0.99 (0.952, 1.032) | 0.6739 | 45,864 | 1.01 (0.993, 1.035) | 0.2053 |
| 4 weeks | 41,695 | 0.99 (0.952, 1.031) | 0.6412 | 45,864 | 1.01 (0.995, 1.024) | 0.1918 |
| **Self-harm** | | | | | | |
|  | **Spring** | | | **Autumn** | | |
|  | **Events** | **IRR (95% CI)** | **p value** | **Events** | **IRR (95% CI)** | **p value** |
| Sunday | 2812 | 0.94 (0.812, 1.097) | 0.4518 | 2,986 | 1.03 (0.959, 1.096) | 0.4660 |
| Monday | 3137 | 1.11 (1.034, 1.199) | 0.0044 | 3,205 | 0.93 (0.841, 1.020 | 0.1210 |
| Tuesday | 3012 | 0.96 (0.868, 1.063) | 0.4403 | 2,938 | 1.00 (0.865, 1.163 | 0.9643 |
| Wednesday | 3022 | 1.07 (0.932, 1.221) | 0.3486 | 2,934 | 0.94 (0.862, 1.015) | 0.1084 |
| Thursday | 2969 | 1.01 (0.958, 1.056) | 0.8150 | 2,813 | 0.98 (0.906, 1.064 | 0.6549 |
| Friday | 2869 | 1.07 (0.942, 1.210) | 0.3036 | 2,854 | 0.97 (0.846, 1.116) | 0.6832 |
| Saturday | 2679 | 0.98 (0.820, 1.165) | 0.7975 | 2,684 | 0.99 (0.862, 1.137) | 0.8887 |
| Mon-Fri | 15,009 | 1.04 (1.004, 1.088) | 0.0332 | 14,744 | 0.96 (0.907, 1.023) | 0.2186 |
| 1 week | 20,500 | 1.02 (0.981, 1.065) | 0.2872 | 20,414 | 0.98 (0.929, 1.023) | 0.3022 |
| 2 weeks | 20,500 | 1.01 (0.990, 1.038) | 0.2719 | 20,414 | 1.00 (0.966, 1.026) | 0.7788 |
| 4 weeks | 20,500 | 0.99 (0.968, 1.007) | 0.2111 | 20,414 | 0.97 (0.938, 1.005) | 0.0887 |
| **Sleep disorders** | | | | | | |
|  | **Spring** | | | **Autumn** | | |
|  | **Events** | **IRR (95% CI)** | **p value** | **Events** | **IRR (95% CI)** | **p value** |
| Sunday | 123 | 0.91 (0.364, 2.289 | 0.8465 | 154 | 1.35 (0.915, 1.981) | 0.1311 |
| Monday | 6891 | 0.94 (0.868, 1.016 | 0.1167 | 7,273 | 0.94 (0.849, 1.041) | 0.2345 |
| Tuesday | 6702 | 1.01 (0.913, 1.118 | 0.8404 | 6,419 | 0.87 (0.781, 0.966) | 0.0095 |
| Wednesday | 6277 | 0.93 (0.842, 1.018 | 0.1113 | 6,106 | 0.92 (0.828, 1.024) | 0.1289 |
| Thursday | 6101 | 1.04 (0.962, 1.127) | 0.3197 | 5,878 | 0.90 (0.831, 0.983) | 0.0181 |
| Friday | 5468 | 1.07 (0.962, 1.187 | 0.2189 | 6,042 | 0.95 (0.885, 1.020) | 0.1540 |
| Saturday | 282 | 0.98 (0.738, 1.290 | 0.8633 | 274 | 0.73 (0.500, 1.076) | 0.1131 |
| Mon-Fri | 31,439 | 1.00 (0.967, 1.034) | 0.9997 | 31,748 | 0.92 (0.869, 0.967) | 0.0013 |
| 1 week | 31,844 | 1.00 (0.965, 1.032) | 0.9082 | 32,146 | 0.92 (0.870, 0.969) | 0.0019 |
| 2 weeks | 31,844 | 0.98 (0.954, 1.009) | 0.1749 | 32,146 | 0.93 (0.895, 0.963) | 0.0001 |
| 4 weeks | 31,844 | 0.96 (0.918, 0.997) | 0.0350 | 32,146 | 0.97 (0.938, 0.995) | 0.0213 |

IRR = Incidence Rate Ratio resulting from negative binomial regression. CI = Confidence Interval.

Events = total number of events in 8-week period (4 weeks before & 4 weeks after the clock change).

Data split by year (12 years), region (9 regions) & day (56 days in 8 week period) which gave us 6,048 data points for the 1,2 and 4 week analyses, 4,320 data points for the Monday-Friday analyses and 864 data points for the individual day analyses.

*Adjustments:

1 week, 2 weeks, 4 weeks & Monday-Friday analyses adjusted for day of the week and region. In addition, Spring analyses were adjusted for the 5 days of the Easter weekend.

Individual day analyses were adjusted for region.

Spring individual day analyses also adjusted for the 5 days of the Easter weekend (except for the Tuesday & Wednesday after where the Easter weekend covariate was omitted from analyses because of collinearity).

Number of events adjusted for the Sunday of the clock change having 25 or 23 hours instead of 24 using the following formulas: Spring: (events on Sunday of clock change / 23) x 24. Autumn: (events on Sunday of clock change / 25) x 24.

**Control periods defined as:

- 1 week after clock change analysis: 4 weeks before & weeks 2-4 after the clock change.

- 2 weeks after clock change analysis: 4 weeks before & weeks 3-4 after the clock change.

- 4 weeks after clock change analysis: 4 weeks before the clock change.

- Monday-Friday in the week after the clock change analysis: weekdays in the 4 weeks before the clock change & weekdays in weeks 2-4 after the clock change.

### Table S5: Sensitivity analysis: Adjusted* rate ratios of the number of health events in the week after the Spring and Autumn clock changes (England, 2008-2019) versus control period**, with events for people with 20 or more events for the same health condition in a single 8-week clock change period excluded from that period & one random event kept for the remaining patients

|  | **Spring (clocks go forward – less sleep)** | | | **Autumn (clocks go back – more sleep)** | | |
| --- | --- | --- | --- | --- | --- | --- |
|  | **Events** | **IRR (95% CI)** | **P value** | **Events** | **IRR (95% CI)** | **P value** |
| Anxiety | 75,740 | 0.98 ( 0.942,1.014) | 0.2165 | 82,563 | 0.95 ( 0.936,0.972) | 7.204x10^-07^ |
| Acute CVD | 188,734 | 1.00 (0.990,1.018) | 0.5819 | 191,512 | 0.97 ( 0.951,0.982) | 3.838x10^-05^ |
| Depression | 209,184 | 0.99 (0.965,1.005) | 0.1496 | 217,498 | 0.95 (0.938,0.963) | 6.826x10^-14^ |
| Eating disorders | 2,085 | 0.98 (0.889,1.076) | 0.6458 | 2,146 | 0.99 (0.864,1.123) | 0.8227 |
| Psychiatric conditions in A&E | 17,293 | 0.99 (0.936,1.040) | 0.6165 | 18,867 | 0.94 (0.898,0.985) | 0.0088 |
| Road traffic injuries | 36,440 | 0.99 (0.946,1.033) | 0.5940 | 39,945 | 0.96 (0.931,0.989) | 0.0069 |
| Self-harm | 17,170 | 1.01 (0.966,1.066) | 0.5621 | 17,054 | 0.96 (0.903,1.021) | 0.1921 |
| Sleep disorders | 28,728 | 0.99 (0.951,1.023) | 0.4747 | 28,917 | 0.91 (0.855,0.962) | 0.0011 |

IRR = Incidence Rate Ratio resulting from negative binomial regression. CI = Confidence Interval.

Events = total number of events in 8-week period (4 weeks before & 4 weeks after the clock change).

Data split by year (12 years), region (9 regions) & day (56 days in 8 week period) which gave us 6,048 data points.

*Analyses adjusted for day of week and region. In addition, Spring analyses were adjusted for the 5 days of the Easter weekend.

**Control period: 4 weeks before & weeks 2-4 after the clock change.

Number of events adjusted for the Sunday of the clock change having 25 or 23 hours instead of 24 using the following formulas: Spring: (events on Sunday of clock change / 23) x 24. Autumn: (events on Sunday of clock change / 25) x 24.

### Table S6: Sensitivity analysis: Adjusted* rate ratios of the number of health events in the week after the Spring and Autumn clock changes (England, 2008-2019) versus control period**, with events of patients with an event for the same health condition in the 8 weeks before the study period removed for that clock change and only the first event for that health condition in the 8 week period kept for the remaining patients

|  | **Spring (clocks go forward – less sleep)** | | | **Autumn (clocks go back – more sleep)** | | |
| --- | --- | --- | --- | --- | --- | --- |
|  | **Events** | **IRR (95% CI)** | **P value** | **Events** | **IRR (95% CI)** | **P value** |
| Anxiety | 63,278 | 0.98 (0.939,1.020) | 0.3145 | 69,691 | 0.96 (0.940,0.988) | 0.0035 |
| Acute CVD | 155,442 | 1.01 (0.999,1.028) | 0.0637 | 159,356 | 0.97 (0.953,0.986) | 0.0003 |
| Depression | 168,097 | 0.98 (0.957,0.997) | 0.0241 | 178,174 | 0.95 (0.934,0.968) | 1.713x10^-08^ |
| Eating disorders | 1,694 | 1.00 (0.943,1.058) | 0.9664 | 1,747 | 0.98 (0.857,1.112) | 0.7180 |
| Psychiatric conditions in A&E | 15,937 | 0.99 (0.930,1.055) | 0.7654 | 17,487 | 0.94 (0.895,0.994) | 0.0283 |
| Road traffic injuries | 35,565 | 0.99 (0.945,1.036) | 0.6512 | 39,015 | 0.97 (0.944,1.005) | 0.1027 |
| Self-harm | 15,465 | 1.03 (0.981,1.074) | 0.2507 | 15,221 | 0.97 (0.904,1.042) | 0.4119 |
| Sleep disorders | 25,042 | 0.99 (0.952,1.024) | 0.4915 | 25,754 | 0.88 (0.827,0.939) | 0.0001 |

IRR = Incidence Rate Ratio resulting from negative binomial regression. CI = Confidence Interval.

Events = total number of events in 8-week period (4 weeks before & 4 weeks after the clock change). Data split by year (12 years), region (9 regions) & day (56 days in 8 week period) which gave us 6,048 data points.

*Analyses adjusted for day of week and region. In addition, Spring analyses were adjusted for the 5 days of the Easter weekend.

**Control period: 4 weeks before & weeks 2-4 after the negative control date.

Number of events adjusted for the Sunday of the clock change having 25 or 23 hours instead of 24 using the following formulas: Spring: (events on Sunday of clock change / 23) x 24. Autumn: (events on Sunday of clock change / 25) x 24.

### Table S7: Secondary analysis: Adjusted rate ratios of the number of health events in the week after the Spring and Autumn clock changes compared to the control period (England, 2008-2019), stratified by sociodemographics

| **ANXIETY** | | | | | | | | |
| --- | --- | --- | --- | --- | --- | --- | --- | --- |
|  | **Spring** | | | | **Autumn** | | | |
|  | **Events** | **IRR (95% CI)** | **P value** | **Cochran’s Q p value** | **Events** | **IRR (95% CI)** | **P value** | **Cochran’s Q**  **p value** |
| **Sex** | | | | | | | | |
| Male | 30859 | 1.01 (0.995,1.035) | 0.1501 | 0.218 | 33379 | 0.96 (0.945,0.972) | 4.604x10^-09^ | 0.548 |
| Female | 64065 | 0.98 (0.938,1.030) | 0.4749 |  | 70632 | 0.97 (0.940,0.998) | 0.0367 |  |
| **Age** | | | | | | | | |
| 10-50 | 50938 | 1.00 (0.958,1.034) | 0.8079 | 0.997 | 55539 | 0.95 (0.917,0.974) | 0.0002 | 0.011 |
| >50 | 43985 | 1.00 (0.950,1.042) | 0.8386 |  | 48474 | 0.99 (0.972,1.003) | 0.1167 |  |
| **Deprivation** | | | | | | | | |
| Most deprived | 27058 | 0.99 (0.938,1.041) | 0.6444 | 0.732 | 30244 | 0.96 (0.943,0.984) | 0.0006 | 0.763 |
| Rest | 67745 | 1.00 (0.966,1.033) | 0.9326 |  | 73663 | 0.97 (0.947,0.989) | 0.0030 |  |
| **Alcohol Status** | | | | | | | | |
| Current drinker | 54801 | 0.99 (0.951,1.037) | 0.7454 | 0.930 | 59695 | 0.98 (0.959,1.001) | 0.0651 | 0.036 |
| Non/ex-drinker | 23390 | 0.99 (0.950,1.038) | 0.7583 |  | 25437 | 0.96 (0.919,1.012) | 0.1428 |  |
| Missing | 16730 | 1.01 (0.936,1.088) | 0.8186 |  | 18885 | 0.92 (0.884,0.961) | 0.0001 |  |
| **Incident case** | | | | | | | | |
| Yes | 31102 | 0.99 (0.936,1.047) | 0.7220 | 0.800 | 34625 | 0.97 (0.933,1.016) | 0.2131 | 0.517 |
| No | 63822 | 1.00 (0.965,1.033) | 0.9251 |  | 69390 | 0.96 (0.939,0.978) | 3.132x10^-05^ |  |
| **CARDIOVASCULAR DISEASE (ACUTE)** | | | | | | | | |
|  | **Spring** | | | | **Autumn** | | | |
|  | **Events** | **IRR (95% CI)** | **P value** | **Cochran’s Q p value** | **Events** | **IRR (95% CI)** | **P value** | **Cochran’s Q**  **p value** |
| **Sex** | | | | | | | | |
| Male | 160675 | 1.02 (1.009,1.040) | 0.0018 | 0.373 | 161329 | 0.97 (0.945,1.006) | 0.1071 | 0.756 |
| Female | 138130 | 1.01 (0.988,1.036) | 0.3450 |  | 140163 | 0.98 (0.957,1.006) | 0.1288 |  |
| **Age** | | | | | | | | |
| 40-75 | 139136 | 1.03 (1.012,1.042) | 0.0003 | 0.163 | 138733 | 0.96 (0.932,0.993) | 0.0179 | 0.095 |
| >75 | 159660 | 1.01 (0.986,1.031) | 0.4792 |  | 162766 | 0.99 (0.972,1.017) | 0.6176 |  |
| **Deprivation** | | | | | | | | |
| Most deprived | 67465 | 1.02 (1.006,1.044) | 0.0078 | 0.403 | 67754 | 1.00 (0.974,1.027) | 0.9802 | 0.122 |
| Rest | 231110 | 1.01 (0.997,1.032) | 0.1165 |  | 233570 | 0.97 (0.949,0.996) | 0.0236 |  |
| **BMI** | | | | | | | | |
| Overweight/obese | 180424 | 1.03 (1.020,1.044) | 2.499x10^-07^ | 0.010 | 183539 | 0.99 (0.963,1.009) | 0.2148 | 0.568 |
| Normal/underweight | 88709 | 1.00 (0.972,1.018) | 0.6793 |  | 89096 | 0.97 (0.939,1.008) | 0.1282 |  |
| Missing | 29655 | 1.00 (0.956,1.040) | 0.8914 |  | 28886 | 0.95 (0.901,1.012) | 0.1194 |  |
| **Alcohol Status** | | | | | | | | |
| Current drinker | 188075 | 1.02 (1.005,1.040) | 0.0119 | 0.382 | 190168 | 0.98 (0.948,1.005) | 0.1017 | 0.742 |
| Non/ex-drinker | 78198 | 1.02 (1.003,1.036) | 0.0175 |  | 79467 | 0.99 (0.959,1.016) | 0.3931 |  |
| Missing | 32514 | 0.98 (0.932,1.037) | 0.5311 |  | 31887 | 0.97 (0.938,1.005) | 0.0933 |  |
| **Smoking Status** | | | | | | | | |
| Current smoker | 39240 | 1.01 (0.991,1.039) | 0.2299 | 0.867 | 40038 | 0.97 (0.935,0.999) | 0.0408 | 0.745 |
| Non/ex-smoker | 247418 | 1.02 (1.002,1.033) | 0.0252 |  | 249503 | 0.98 (0.957,1.007) | 0.1603 |  |
| Missing | 12125 | 1.00 (0.938,1.066) | 0.9920 |  | 11987 | 0.97 (0.928,1.020) | 0.2529 |  |
| **Systolic Blood Pressure** | | | | | | | | |
| High | 101233 | 1.03 (1.003,1.058) | 0.0277 | 0.314 | 101206 | 0.98 (0.950,1.008) | 0.1441 | 0.450 |
| Normal | 185045 | 1.01 (0.998,1.021) | 0.1107 |  | 188033 | 0.98 (0.964,1.001) | 0.0600 |  |
| Missing | 12510 | 1.03 (0.974,1.093) | 0.2853 |  | 12280 | 0.94 (0.888,1.003) | 0.0608 |  |
| **Diastolic Blood Pressure** | | | | | | | | |
| High | 26606 | 1.01 (0.972,1.056) | 0.5493 | 0.874 | 27596 | 0.96 (0.924,1.006) | 0.0957 | 0.391 |
| Normal | 259669 | 1.02 (1.003,1.031) | 0.0174 |  | 261647 | 0.98 (0.962,1.003) | 0.0873 |  |
| Missing | 12512 | 1.03 (0.974,1.093) | 0.2884 |  | 12281 | 0.94 (0.888,1.003) | 0.0603 |  |
| **Incident case** | | | | | | | | |
| Yes | 58511 | 1.02 (0.999,1.048) | 0.0648 | 0.814 | 58905 | 0.99 (0.964,1.016) | 0.4531 | 0.472 |
| No | 240279 | 1.02 (1.006,1.033) | 0.0055 |  | 242617 | 0.98 (0.956,0.999) | 0.0450 |  |
| **CVD subgroup (GP & HES APC data only)** | | | | | | | | |
| Atrial Fibrillation / Arrhythmia | 107497 | 1.03 (1.009,1.042) | 0.0021 | 0.654 | 107146 | 0.98 (0.958,1.008) | 0.1872 | 0.698 |
| Myocardial Infarction | 11245 | 1.00 (0.950,1.050) | 0.9510 |  | 11044 | 0.96 (0.916,1.016) | 0.1729 |  |
| Stroke / TIA | 35788 | 1.03 (0.983,1.085) | 0.1975 |  | 35014 | 1.00 (0.947,1.058) | 0.9708 |  |
| Other | 80910 | 1.01 (0.993,1.036) | 0.1881 |  | 81798 | 0.97 (0.947,0.995) | 0.0184 |  |
| **DEPRESSION** | | | | | | | | |
|  | **Spring** | | | | **Autumn** | | | |
|  | **Events** | **IRR (95% CI)** | **P value** | **Cochran’s Q p value** | **Events** | **IRR (95% CI)** | **P value** | **Cochran’s Q**  **p value** |
| **Sex** |  |  |  |  |  |  |  |  |
| Male | 84687 | 1.00 (0.985,1.022) | 0.7021 | 0.266 | 88845 | 0.96 **(**0.947,0.975) | 7.359x10^-08^ | 0.711 |
| Female | 173723 | 0.99 (0.974,1.006) | 0.2059 |  | 180221 | 0.96 (0.940,0.974) | 1.202x10^-06^ |  |
| **Age** | | | | | | | | |
| 10-45 | 127747 | 0.99 (0.970,1.013) | 0.4439 | 0.771 | 131121 | 0.94 (0.915,0.958) | 1.736x10^-08^ | 0.015 |
| >45 | 130670 | 1.00 (0.980,1.012) | 0.5885 |  | 137958 | 0.98 (0.952,1.004) | 0.0927 |  |
| **Deprivation** | | | | | | | | |
| Most deprived | 76732 | 0.98 (0.966,0.990) | 0.0002 | 0.027 | 79666 | 0.96 (0.937,0.988) | 0.0037 | 0.647 |
| Rest | 181392 | 1.00 (0.984,1.024) | 0.7152 |  | 189162 | 0.96 (0.949,0.963) | 1.565x10^-33^ |  |
| **Alcohol Status** | | | | | | | | |
| Current drinker | 157444 | 0.99 (0.972,1.010) | 0.3628 | 0.977 | 163854 | 0.96 (0.941,0.978) | 3.031x10^-05^ | 0.528 |
| Non/ex-drinker | 57519 | 0.99 (0.970,1.020) | 0.6690 |  | 59415 | 0.96 (0.943,0.982) | 0.0002 |  |
| Missing | 43450 | 0.99 (0.962,1.025) | 0.6636 |  | 45817 | 0.95 (0.923,0.969) | 5.939x10^-06^ |  |
| **Incident case** | | | | | | | | |
| Yes | 51392 | 0.98 (0.958,1.006) | 0.1310 | 0.218 | 54133 | 0.98 (0.960,0.997) | 0.0217 | 0.032 |
| No | 207037 | 1.00 (0.983,1.017) | 0.9874 |  | 214940 | 0.95 (0.938,0.967) | 5.110x10^-10^ |  |
| **EATING DISORDERS** | | | | | | | | |
|  | **Spring** | | | | **Autumn** | | | |
|  | **Events** | **IRR (95% CI)** | **P value** | **Cochran’s Q p value** | **Events** | **IRR (95% CI)** | **P value** | **Cochran’s Q**  **p value** |
| **Sex** |  |  |  |  |  |  |  |  |
| Male | 211 | 1.40 (0.927,2.108) | 0.1100 | 0.164 | 208 | 1.05 (0.755,1.466) | 0.7657 | 0.655 |
| Female | 2524 | 0.97 (0.846,1.108) | 0.6422 |  | 2546 | 0.97 (0.883,1.062) | 0.4915 |  |
| **Age** |  |  |  |  |  |  |  |  |
| 10-25 | 1295 | 1.02 (0.930,1.123) | 0.6482 | 0.618 | 1316 | 0.99 (0.856,1.143) | 0.8869 | 0.763 |
| >25 | 1440 | 0.98 (0.861,1.118) | 0.7761 |  | 1438 | 0.96 (0.856,1.080) | 0.5087 |  |
| **Deprivation** | | | | | | | | |
| Most deprived | 561 | 1.04 (0.784,1.378) | 0.7877 | 0.816 | 620 | 1.17 (0.840,1.642) | 0.3481 | 0.220 |
| Rest | 2158 | 1.00 (0.920,1.093) | 0.9498 |  | 2130 | 0.92 (0.841,1.003) | 0.0572 |  |
| **BMI** | | | | | | | | |
| Overweight/obese | 319 | 1.02 (0.806,1.302) | 0.8433 | 0.879 | 310 | 0.98 (0.656,1.458) | 0.9139 | 0.988 |
| Normal/underweight | 1668 | 1.01 (0.898,1.137) | 0.8647 |  | 1606 | 0.97 (0.884,1.062) | 0.4967 |  |
| Missing | 748 | 0.97 (0.868,1.092) | 0.6461 |  | 838 | 0.98 (0.831,1.164) | 0.8451 |  |
| **Incident case** | | | | | | | | |
| Yes | 845 | 0.92 (0.770,1.088) | 0.3148 | 0.209 | 830 | 1.11 (0.869,1.418) | 0.4016 | 0.198 |
| No | 1890 | 1.04 (0.936,1.152) | 0.4770 |  | 1924 | 0.92 (0.825,1.024) | 0.1247 |  |
| **PSYCHIATRIC CONDITIONS (IN A&E ONLY)** | | | | | | | | |
|  | **Spring** | | | | **Autumn** | | | |
|  | **Events** | **IRR (95% CI)** | **P value** | **Cochran’s Q p value** | **Events** | **IRR (95% CI)** | **P value** | **Cochran’s Q**  **p value** |
| **Sex** |  |  |  |  |  |  |  |  |
| Male | 9289 | 0.98 (0.895,1.064) | 0.5814 | 0.813 | 10170 | 1.00 (0.948,1.057) | 0.9589 | 0.005 |
| Female | 9976 | 0.99 (0.934,1.045) | 0.6755 |  | 10906 | 0.89 (0.835,0.947) | 0.0003 |  |
| **Age** |  |  |  |  |  |  |  |  |
| 10-35 | 9430 | 0.97 (0.891,1.059) | 0.5065 | 0.704 | 10099 | 0.90 (0.843,0.954) | 0.0006 | 0.029 |
| >35 | 9835 | 0.99 (0.924,1.066) | 0.8359 |  | 10977 | 0.99 (0.930,1.044) | 0.6226 |  |
| **Deprivation** | | | | | | | | |
| Most deprived | 7416 | 0.97 (0.877,1.066) | 0.4958 | 0.718 | 8044 | 0.93 (0.876,0.989) | 0.0199 | 0.673 |
| Rest | 11828 | 0.99 (0.921,1.060) | 0.7438 |  | 13001 | 0.95 (0.880,1.027) | 0.1990 |  |
| **Alcohol Status** | | | | | | | | |
| Current drinker | 8541 | 1.02 (0.929,1.126) | 0.6493 | 0.389 | 9137 | 0.95 (0.902,0.992) | 0.0225 | 0.002 |
| Non/ex-drinker | 4134 | 0.93 (0.857,1.019) | 0.1236 |  | 4600 | 1.05 (0.987,1.119) | 0.1193 |  |
| Missing | 6590 | 0.96 (0.912,1.011) | 0.1232 |  | 7339 | 0.87 (0.800,0.957) | 0.0034 |  |
| **Incident case** | | | | | | | | |
| Yes | 11844 | 1.03 (0.988,1.076) | 0.1649 | 0.019 | 13000 | 0.92 (0.858,0.984) | 0.0151 | 0.129 |
| No | 7421 | 0.90 (0.810,1.006) | 0.0631 |  | 8076 | 0.99 (0.927,1.052) | 0.6967 |  |
| **ROAD TRAFFIC INJURIES** | | | | | | | | |
|  | **Spring** | | | | **Autumn** | | | |
|  | **Events** | **IRR (95% CI)** | **P value** | **Cochran’s Q p value** | **Events** | **IRR (95% CI)** | **P value** | **Cochran’s Q**  **p value** |
| **Sex** |  |  |  |  |  |  |  |  |
| Male | 22798 | 0.97 (0.922,1.026) | 0.3124 | 0.400 | 25097 | 0.98 (0.946,1.024) | 0.4351 | 0.539 |
| Female | 18889 | 1.01 (0.944,1.080) | 0.7819 |  | 20773 | 0.97 (0.926,1.009) | 0.1218 |  |
| **Age** |  |  |  |  |  |  |  |  |
| 0-30 | 19024 | 1.02 (0.956,1.079) | 0.6108 | 0.248 | 20586 | 0.99 (0.937,1.048) | 0.7441 | 0.469 |
| >30 | 22665 | 0.97 (0.918,1.022) | 0.2395 |  | 25286 | 0.97 (0.928,1.005) | 0.0884 |  |
| **Deprivation** | | | | | | | | |
| Most deprived | 12327 | 0.96 (0.892,1.028) | 0.2336 | 0.279 | 13179 | 0.93 (0.857,1.015) | 0.1054 | 0.163 |
| Rest | 29333 | 1.00 (0.960,1.046) | 0.9218 |  | 32657 | 0.99 (0.961,1.027) | 0.6897 |  |
| **Alcohol Status** | | | | | | | | |
| Current drinker | 19015 | 0.95 (0.911,1.000) | 0.0498 | 0.038 | 21229 | 0.96 (0.935,0.994) | 0.0172 | 0.002 |
| Non/ex-drinker | 6956 | 0.95 (0.892,1.011) | 0.1070 |  | 7477 | 0.89 (0.851,0.938) | 6.466x10^-06^ |  |
| Missing | 15717 | 1.05 (0.986,1.116) | 0.1284 |  | 17166 | 1.03 (0.965,1.100) | 0.3692 |  |
| **SELF-HARM** | | | | | | | | |
|  | **Spring** | | | | **Autumn** | | | |
|  | **Events** | **IRR (95% CI)** | **P value** | **Cochran’s Q p value** | **Events** | **IRR (95% CI)** | **P value** | **Cochran’s Q**  **p value** |
| **Sex** |  |  |  |  |  |  |  |  |
| Male | 8438 | 1.04 (0.964,1.112) | 0.3432 | 0.592 | 8551 | 1.02 (0.946,1.102) | 0.5961 | 0.152 |
| Female | 12056 | 1.01 (0.968,1.057) | 0.6091 |  | 11864 | 0.95 (0.882,1.015) | 0.1211 |  |
| **Age** | | | | | | | | |
| 10-30 | 10311 | 1.03 (0.965,1.089) | 0.4204 | **0.838** | 9962 | 0.95 (0.893,1.009) | 0.0928 | 0.288 |
| >30 | 10187 | 1.02 (0.979,1.058) | 0.3759 |  | 10455 | 1.00 (0.927,1.083) | 0.9595 |  |
| **Deprivation** | | | | | | | | |
| Most deprived | 7692 | 1.08 (1.023,1.141) | 0.0054 | 0.036 | 7559 | 0.94 (0.865,1.026) | 0.1734 | 0.324 |
| Rest | 12771 | 0.99 (0.931,1.052) | 0.7409 |  | 12814 | 0.99 (0.934,1.056) | 0.8240 |  |
| **Alcohol Status** | | | | | | | | |
| Current drinker | 8524 | 0.99 (0.938,1.049) | 0.7807 | 0.308 | 8687 | 1.02 (0.973,1.079) | 0.3648 | 0.117 |
| Non/ex-drinker | 3523 | 1.06 (0.989,1.146) | 0.0978 |  | 3587 | 0.93 (0.827,1.036) | 0.1809 |  |
| Missing | 8451 | 1.03 (0.968,1.104) | 0.3160 |  | 8143 | 0.95 (0.881,1.023) | 0.1716 |  |
| **Incident case** | | | | | | | | |
| Yes | 9893 | 1.02 (0.961,1.083) | 0.5083 | 0.937 | 9526 | 0.92 (0.890,0.960) | 4.082x10^-05^ | 0.018 |
| No | 10605 | 1.02 (0.965,1.086) | 0.4334 |  | 10891 | 1.02 (0.951,1.099) | 0.5486 |  |
| **SLEEP** | | | | | | | | |
|  | **Spring** | | | | **Autumn** | | | |
|  | **Events** | **IRR (95% CI)** | **P value** | **Cochran’s Q**  **p value** | **Events** | **IRR (95% CI)** | **P value** | **Cochran’s Q**  **p value** |
| **Sex** |  |  |  |  |  |  |  |  |
| Male | 12651 | 0.99 (0.957,1.021) | 0.4871 | 0.563 | 12711 | 0.92 (0.856,0.992) | 0.0297 | 0.925 |
| Female | 19192 | 1.01 (0.955,1.061) | 0.8006 |  | 19434 | 0.93 (0.861,0.996) | 0.0375 |  |
| **Age** |  |  |  |  |  |  |  |  |
| 10-55 | 16684 | 1.00 (0.971,1.024) | 0.8367 | 0.857 | 16784 | 0.94 (0.891,0.982) | 0.0066 | 0.415 |
| >55 | 15160 | 1.00 (0.958,1.049) | 0.9292 |  | 15362 | 0.91 (0.866,0.954) | 0.0001 |  |
| **Deprivation** | | | | | | | | |
| Most deprived | 7593 | 1.00 (0.935,1.080) | 0.9016 | 0.973 | 7646 | 0.95 (0.897,1.006) | 0.0767 | 0.284 |
| Rest | 24222 | 1.00 (0.969,1.038) | 0.8578 |  | 24480 | 0.91 (0.852,0.965) | 0.0021 |  |
| **Alcohol Status** | | | | | | | | |
| Current drinker | 20779 | 1.01 (0.974,1.049) | 0.5756 | 0.258 | 20876 | 0.90 (0.841,0.970) | 0.0051 | 0.341 |
| Non/ex-drinker | 6972 | 0.96 (0.909,1.012) | 0.1273 |  | 7162 | 0.97 (0.912,1.025) | 0.2610 |  |
| Missing | 4093 | 1.01 (0.928,1.109) | 0.7541 |  | 4108 | 0.94 (0.895,0.993) | 0.0255 |  |
| **Incident case** | | | | | | | | |
| Yes | 14670 | 1.03 (0.978,1.081) | 0.2764 | 0.123 | 15204 | 0.90 (0.851,0.961) | 0.0012 | 0.352 |
| No | 17174 | 0.97 (0.929,1.021) | 0.2726 |  | 16942 | 0.94 (0.894,0.985) | 0.0104 |  |

IRR = Incidence Rate Ratio resulting from negative binomial regression. CI = Confidence Interval.

Events = total number of events in 8-week period (4 weeks before & 4 weeks after the clock change).

Data split by year (12 years), region (9 regions) & day (56 days in 8 week period) which gave us 6,048 data points.

Control period: 4 weeks before & weeks 2-4 after the negative control date.

Analyses adjusted for day of week and region. In addition, Spring analyses were adjusted for the 5 days of the Easter weekend.

Number of events adjusted for the Sunday of the clock change having 25 or 23 hours instead of 24 using the following formulas: Spring: (events on Sunday of clock change / 23) x 24. Autumn: (events on Sunday of clock change / 25) x 24.

### Table S8: Negative control exposure analysis: Adjusted* rate ratios of the number of health events in the week after the Spring and Autumn negative controls (4 weeks before the clock changes) (England, 2008-2019) versus control period**

|  | **Spring negative control** | | | **Autumn negative control** | | |
| --- | --- | --- | --- | --- | --- | --- |
|  | **Events** | **IRR (95% CI)** | **P value** | **Events** | **IRR (95% CI)** | **P value** |
| Anxiety | 96,700 | 1.00 (0.980,1.013) | 0.6749 | 103,150 | 1.01 (0.987,1.032) | 0.4255 |
| Acute CVD | 305,054 | 1.00 (0.986,1.011) | 0.8136 | 296,563 | 1.00 (0.982,1.008) | 0.4507 |
| Depression | 268,746 | 1.01 (0.993,1.023) | 0.3047 | 267,634 | 1.02 (1.006,1.043) | 0.0106 |
| Eating disorders | 2,645 | 0.99 (0.885,1.112) | 0.8862 | 2,686 | 1.02 (0.940,1.101) | 0.6678 |
| Psychiatric conditions in A&E | 19,066 | 1.02 (0.969,1.075) | 0.4438 | 20,846 | 1.00 (0.949,1.061) | 0.9061 |
| Road traffic injuries | 43,293 | 1.01 (0.971,1.041) | 0.7719 | 45,129 | 1.02 (0.977,1.061) | 0.3974 |
| Self-harm | 20,548 | 1.02 (0.986,1.049) | 0.2985 | 20,742 | 1.00 (0.954,1.054) | 0.9120 |
| Sleep disorders | 35,322 | 1.04 (1.021,1.053) | 3.222x10^-06^ | 32,502 | 1.00 (0.962,1.039) | 0.9835 |

IRR = Incidence Rate Ratio resulting from negative binomial regression. CI = Confidence Interval. Events = total number of events in 8-week period (4 weeks before & 4 weeks after the clock change). Data split by year (12 years), region (9 regions) & day (56 days in 8 week period) which gave us 6,048 data points.

*Analyses adjusted for day of week and region. In addition, Spring analyses were adjusted for the 5 days of the Easter weekend.

**Control period: 4 weeks before & weeks 2-4 after the negative control date.

### ****Figure S1: Exposure and control periods for primary and secondary analyses.****

S1a: Primary analysis: Exposed period = first week after the clock change

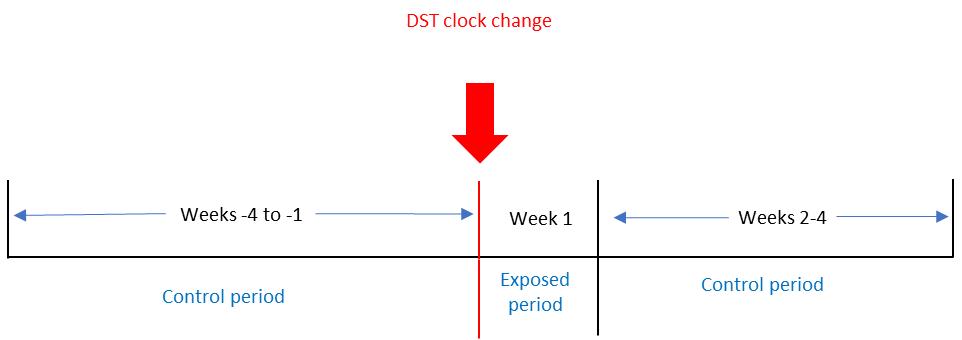

S1b: Secondary analysis. Exposed period = individual days of the week in the first week after the clock change (diagram shows Monday)

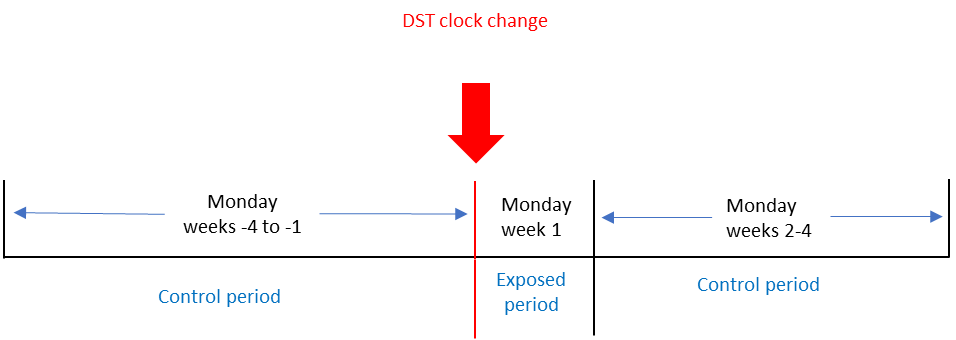

**S1c: Secondary analysis: Exposed period = Monday to Friday in first week after the clock change**

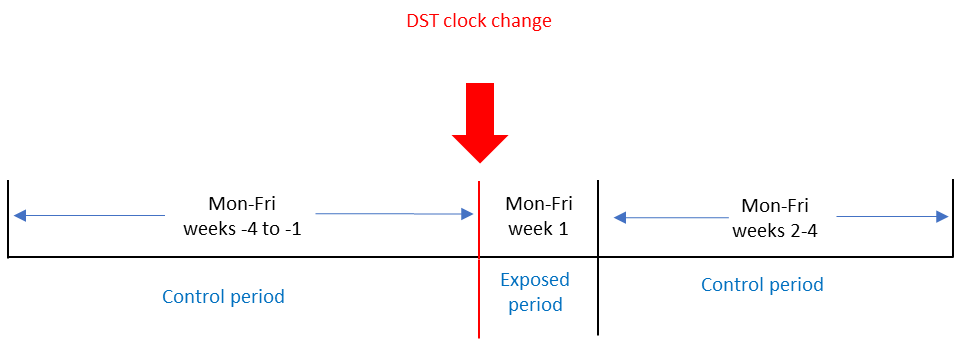

**S1d: Secondary analysis: Exposed period = first 2 weeks after clock change**

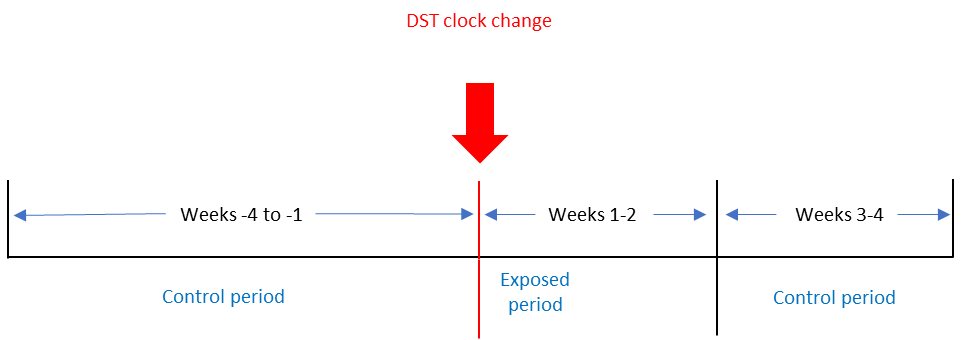

**S1e: Secondary analysis: Exposed period = first 4 weeks after clock change**

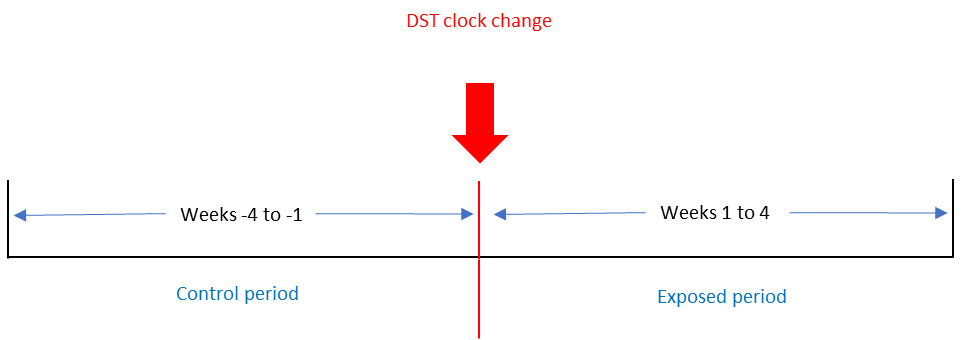

**S1f: Negative control analysis: Exposed period = week 4 before clock change**

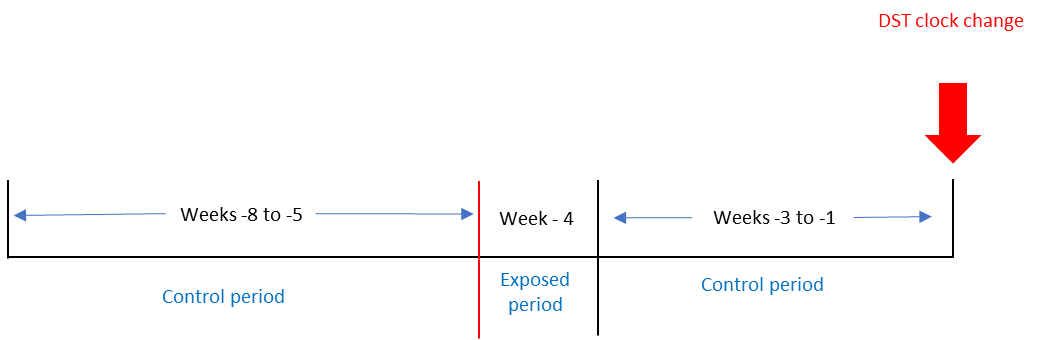

### ****Figure S2: Anxiety events per day over the Spring clock changes.****

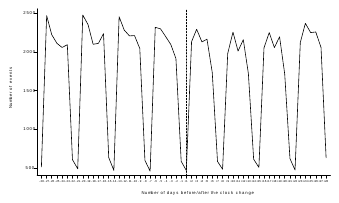

*Dashed line = day of the clock change.

### ****Figure S3: CVD events per day over the Spring clock changes.****

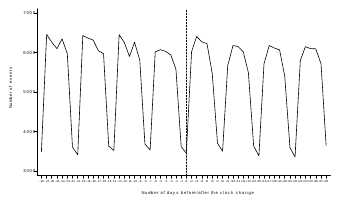

*Dashed line = day of the clock change.

### ****Figure S4: Depression events per day over the Spring clock changes.****

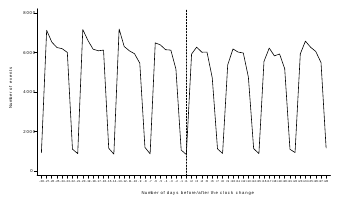

*Dashed line = day of the clock change.

### ****Figure S5: Eating disorder events per day over the Spring clock changes.****

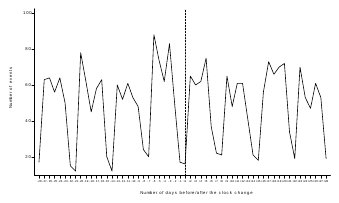

*Dashed line = day of the clock change.

### ****Figure S6: Psychiatric conditions events per day over the Spring clock changes.****

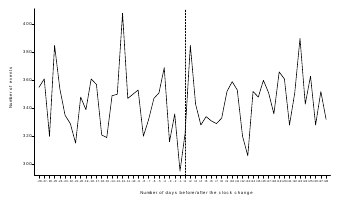

*Dashed line = day of the clock change.

### ****Figure S7: Road traffic injury events per day over the Spring clock changes.****

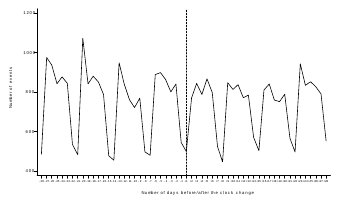

*Dashed line = day of the clock change.

### ****Figure S8: Self-harm events per day over the Spring clock changes.****

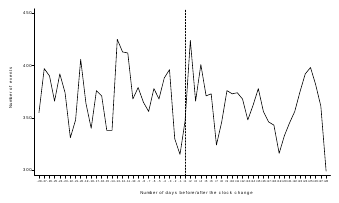

*Dashed line = day of the clock change.

### ****Figure S9: Sleep disorder events per day over the Spring clock changes.****

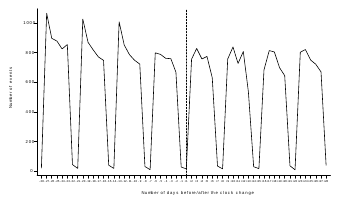

*Dashed line = day of the clock change.

### ****Figure S10: Anxiety events per day over the Autumn clock changes.****

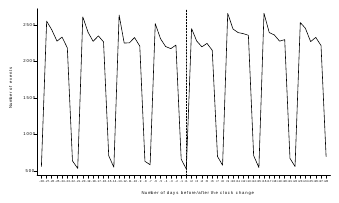

*Dashed line = day of the clock change.

### ****Figure S11: CVD events per day over the Autumn clock changes.****

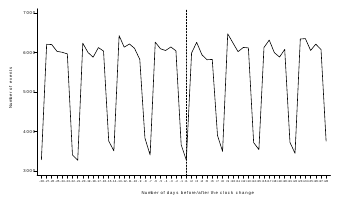

*Dashed line = day of the clock change.

### ****Figure S12: Depression events per day over the Autumn clock changes.****

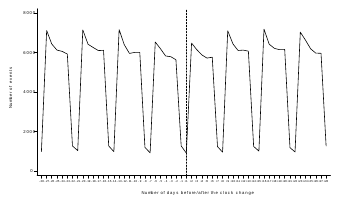

*Dashed line = day of the clock change.

### ****Figure S13: Eating disorder events per day over the Autumn clock changes.****

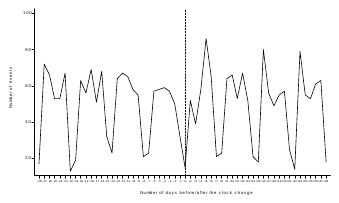

*Dashed line = day of the clock change.

### ****Figure S14: Psychiatric conditions events per day over the Autumn clock changes.****

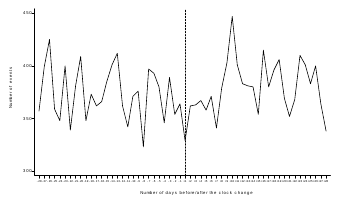

*Dashed line = day of the clock change.

### ****Figure S15: Road traffic injuries events per day over the Autumn clock changes.****

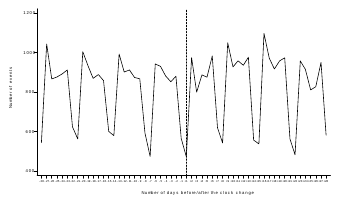

*Dashed line = day of the clock change.

### ****Figure S16: Self-harm events per day over the Autumn clock changes.****

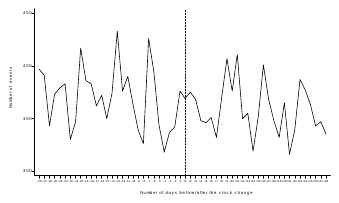

*Dashed line = day of the clock change.

### ****Figure S17: Sleep disorders events per day over the Autumn clock changes.****

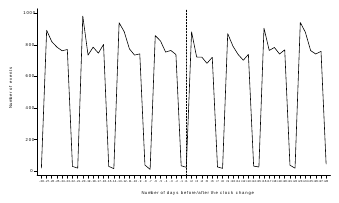

*Dashed line = day of the clock change.
